# Integrating spatial transcriptomics and snRNA-seq data enhances differential gene expression analysis results of AD-related phenotypes

**DOI:** 10.1101/2024.11.18.24317499

**Authors:** Shizhen Tang, Shihan Liu, Aron S. Buchman, David A. Bennett, Philip L. De Jager, Jian Hu, Jingjing Yang

## Abstract

**Background:** Spatial transcriptomics (**ST**) data provide spatially-informed gene expression for studying complex diseases such as Alzheimer’s disease (**AD**). Existing studies using ST data to identify genes with spatially-informed differential gene expression (**DGE**) of complex diseases have limited power due to small sample sizes. Conversely, single-nucleus RNA sequencing (**snRNA-seq**) data offer larger sample sizes for studying cell-type specific (**CTS**) DGE but lack spatial information. In this study, we integrated ST and snRNA-seq data to enhance the power of spatially-informed CTS DGE analysis of AD-related phenotypes.

**Method:** First, we utilized the recently developed deep learning tool **CelEry** to infer the spatial location of ∼1.5M cells from snRNA-seq data profiled from dorsolateral prefrontal cortex (**DLPFC**) tissue of **436** postmortem brains in the ROS/MAP cohorts. Spatial locations of six cortical layers that have distinct anatomical structures and biological functions were inferred. Second, we conducted cortical-layer specific (**CLS**) and CTS DGE analyses for three quantitative AD-related phenotypes –– β-amyloid, tangle density, and cognitive decline. CLS-CTS DGE analyses were conducted based on linear mixed regression models with pseudo-bulk scRNA-seq data and inferred cortical layer locations.

**Results:** We identified 450 potential CLS-CTS significant genes with nominal p-values<10^-4^, including 258 for β-amyloid, 122 for tangle density, and 127 for cognitive decline. Majority of these identified genes, including the ones having known associations with AD (e.g., *APOE*, *KCNIP3*, and *CTSD*), cannot be detected by traditional CTS DGE analyses without considering spatial information. We also identified 8 genes shared across all three phenotypes, 21 between β-amyloid and tangle density, 10 between cognitive decline and tangle density, and 10 between β-amyloid and cognitive density. Particularly, Gene Set Enrichment Analyses with the CLS-CTS DGE results of microglia in cortical layer-6 of β-amyloid identified 12 significant AD-related pathways.

**Conclusion:** Incorporating spatial information with snRNA-seq data detected significant genes and pathways for AD-related phenotypes that would not be identified by traditional CTS DGE analyses. These identified CLS-CTS significant genes not only help illustrate the pathogenesis of AD, but also provide potential CLS-CTS targets for developing therapeutics of AD.

## Background

The technologies of profiling spatial transcriptomics (ST) data have revolutionized our understanding of spatially-informed gene expression, providing invaluable insights into the molecular architectures of complex diseases like Alzheimer’s disease (AD) (1). Despite the additional spatial information, employing ST data to study complex diseases still faces significant challenges. Most spatial transcriptomics studies of the human brain involve small sample sizes (n < 10). As summarized in recent reviews (2), brain tissue datasets predominantly feature ST data from mouse models, with only a few dozens of human samples available. Consequently, human brain data, particularly for studying AD, are significantly limited compared to available single nucleus RNA sequencing (snRNA-seq) datasets. For instance, the recently published HEST-1k dataset (2) summarizes 1,108 ST profiles that are the majority of available public ST data, of which only 210 are of brain samples. The available ST data of brain samples are predominantly profiled from healthy subjects, further limiting the disease-focused research potential. Besides limited sample sizes, another drawback of spatial ST data compared to snRNA-seq data is the trade-off between resolution and gene coverage. Sequencing-based ST can measure the whole genome but only at bulk resolution, while in situ-based ST techniques can measure fewer than 500 genes at single-cell resolution. These limitations severely restrict the potential for conducting spatially-informed and cell type specific (CTS) differential gene expression (DGE) analyses that are critical for studying complex diseases.

In contrast to ST, the technology of snRNA-seq can profile gene expression of transcriptome-wide genes with lower cost, and is widely used for cohorts with larger sample sizes (3,4). Although snRNA-seq data allow CTS DGE analyses, snRNA-seq data lack the spatial contexts of all single nucleus/cells that are critical for fully understanding the cellular and molecular complexity of AD. The spatial arrangement of cells within tissues can reveal spatial specific interactions and functional connectivity that are vital for deciphering the pathogenesis of AD (5).

To detect genes with spatially-informed CTS differential expressions in AD, we integrated public reference ST data provided by LIBD (6) with snRNA-seq data of a large cohort (n=436) (3,4) to conduct cortical-layer specific (CLS) and CTS DGE analyses (**Fig 1**). The snRNA-Seq data were profiled from the dorsolateral prefrontal cortex (DLPFC) tissue of 436 postmortem brains in the Religious Order Study and the Rush Memory and Aging Project (ROS/MAP) cohorts (3,4). We first utilized a deep learning tool, CelEry (7), to infer the spatial locations of six cortical layers for approximately 1.5 million cells in the ROS/MAP snRNAseq data, taking the LIBD ST data as references (**Fig 1A**). Each of the six annotated cortical layers is characterized by distinct anatomical structures and biological functions. Next, we utilized the linear mixed model (LMM) based testing method as implemented by GEMMA (8) to test differential gene expression for each gene-phenotype pair, with the pseudo-bulk scRNA-seq data of each cell type in each cortical layer (**Fig 1B**). We considered three quantitative AD-related phenotypes –– β-amyloid, tangle density, and cognitive decline.

**Fig 1:**
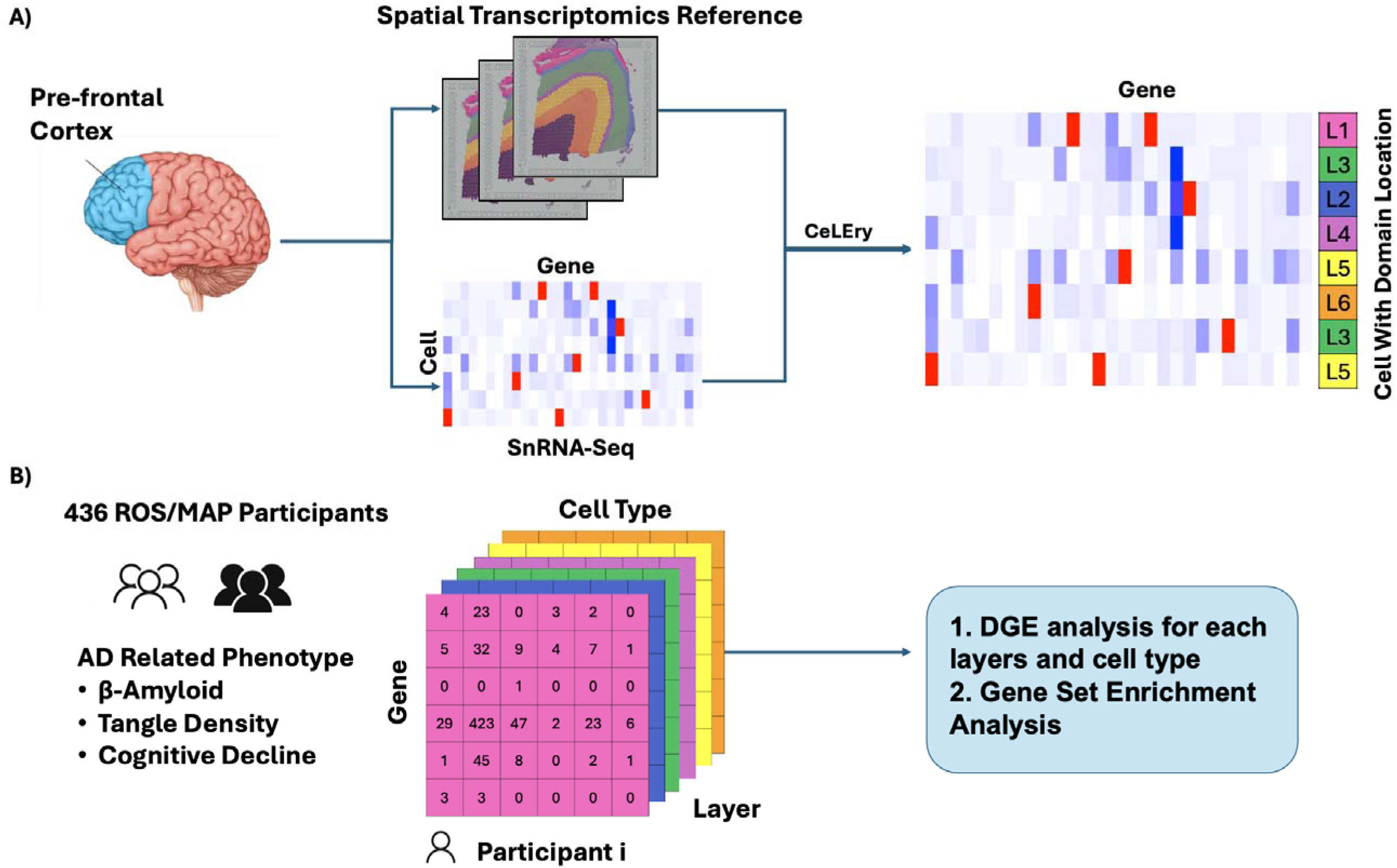
Workflow for CLS-CTS DGE Analyses of AD-related phenotypes. A) CeLEry tool was first used to infer the spatial locations of cells from the ROS/MAP snRNA-Seq data of DLPFC. B) Pseudo bulk read counts were generated from the scRNA-Seq data for each cell type in each cortical layer, which were used to conduct CLS-CTS DGE analyses of three AD-related phenotypes, -amyloid, tangle density, and cognitive decline. Top significant genes from the CLS-CTS DGE analyses were further used to conduct gene set enrichment analyses.

By integrating ST with snRNA-seq data, we identified 450 CLS-CTS significant genes, of which 368 were missed by CTS DGE analyses that does not consider the spatial information. Important AD-related significant pathways were found enriched with the top CLS-CTS significant genes of microglia in cortical layer-6 of β-amyloid. Additionally, we identified important AD related genes that had differential expression across multiple phenotypes only in specific cell types and cortical layers.

In the following sections, we first describe the data sets used in this study, followed by the CeLEry method for annotating spatial information of cells in snRNA-seq data, the LMM model used for DGE analyses, and gene set enrichment analysis procedure in the methods section. Next, we present the results, and discuss the importance of integrating ST and snRNA-seq data for DGE analyses, and end with a conclusion section.

## Methods

### Data preprocessing

#### LIBD dat

The LIBD dataset comprises ST data profiled from three human subjects including two males and one female, each with four replicates, resulting in a total of 12 sections. This dataset contains ST data for 33,538 genes and 47,681 spots, generated by using the Visium platform. For each subject, two spatially adjacent replicates are positioned without gap, with two additional replicates located 300 micrometers away. The cortical layers in this dataset have been manually labeled using the spatialLIBD tool (9), assigning each spot to six cortical layers (L1 to L6) and white matter (WM) (10). L1, known as molecular layer, consists of fiber and synapse-rich regions with few cell bodies, serving as a pathway for numerous parallel axons. L2, external granular layer, mainly contains small pyramidal neurons with apical dendrites directed toward the brain’s surface, playing key roles in cortical connectivity. L3, external pyramidal layer, features a thick arrangement of medium-sized pyramidal neurons that connect extensively within and across brain hemispheres. L4, internal granular layer, is densely packed with stellate and small pyramidal cells, critical for receiving and processing sensory information. L5, internal pyramidal layer, contains medium to large pyramidal neurons, including Betz cells, which project to subcortical regions influencing motor control. L6, multiform Layer, composes of a diverse mix of neuron types, primarily facilitating communication with the thalamus (10). This LIBD ST dataset was used as a reference for inferring cortical layer locations of cells in the ROS/MAP snRNAseq data, for being profiled from the same DLPFC region.

#### CosMx ST data

The CosMx dataset, characterized by a pre-commercial version of the 6078-plex CosMx Spatial Molecular Imager (SMI), is a single-cell resolution ST dataset profiled from human frontal cortex formalin-fixed, paraffin-embedded (FFPE) tissue. Given its single-cell resolution and its origin from the same DLPFC region as the LIBD data and ROS/MAP snRNAsesq data, we utilized the CosMx data to guide our selection of the best reference ST from 12 samples in the LIBD dataset.

To identify brain regions in the CosMax ST data, we used the unsupervised clustering method SpaGCN (11), to divide the ST brain section into distinct domains that closely reflect cortical layer structures. We then assigned each unidentified domain to one of the six cortical layers or WM by adopting the annotation strategy from the spatialLIBD tool. First, we compiled lists of marker genes for the six cortical layers and WM based on previous studies (5,10). Second, we conducted two-sample t-tests for each marker gene to compare its expression in an unidentified domain against the remaining domains. Each unidentified domain is represented by a vector of t-statistics across marker genes. Third, we calculated the correlations between these t-statistic vectors from unidentified domains and corresponding vectors from cortical layers in the LIBD data. Last, we assigned each unidentified domain to the cortical layer or WM with the highest correlation.

#### ROS/MAP snRNA-Seq data

The ROS/MAP dataset originates from two longitudinal, community-based clinic pathological cohort studies: the ROS and MAP (3,4).

Participants were recruited without known dementia at the time of enrollment and agreed to annual clinical evaluations and brain donation upon death. Each study received approval from the Institutional Review Board of Rush University Medical Center, with all participants signing informed consent forms, Anatomical Gift Act agreements, and repository consents. Collectively referred to as ‘ROS/MAP,’ these harmonized cohorts are designed to be analyzed jointly. In this study, the snRNA-seq data were profiled from 436 postmortem brain specimens of DLPFC tissue from the ROS/MAP cohorts, yielding transcriptomic profiles of 1,546,794 cells. Our analyses focused on six major cell types: astrocytes (Ast), excitatory neurons (Ex), inhibitory neurons (In), microglia (Mic), oligodendrocytes (Oli), and oligodendrocyte precursor cells (OPC).

#### AD-related phenotypes

Our study concentrated on three quantitative AD-related phenotypes –– cognitive decline and two AD neuropathologic changes traits (tangle density and β-amyloid). Cognitive decline was measured as the person-specific rate of change in global cognition, estimated as person-specific random effects over all follow-up periods from a linear mixed model (12,13). Tangle density was determined using molecularly specific immunohistochemistry, represented as the average PHFtau tangle density across two or more 20µm sections from eight brain regions: hippocampus, entorhinal cortex, midfrontal cortex, inferior temporal cortex, angular gyrus, calcarine cortex, anterior cingulate cortex, and superior frontal cortex. Similarly, β-amyloid was quantified as the average percentage of cortical area occupied by β-amyloid protein in adjacent sections from the same eight brain regions. To enhance normality, the values of tangle density and β-amyloid were transformed by taking their square root (14,15).

### Infer cortical layers for ROS/MAP snRNA-seq data by CeLEry

We employed the recently developed tool CeLEry (7) to infer the spatial locations of ∼1.5 million cells from the ROS/MAP snRNA-Seq data. CeLEry is a supervised deep learning tool designed to recover the spatial locations of cells in snRNA-seq data. CeLEry trains a deep learning model to capture the relationship between gene expression and spatial locations by using a reference ST dataset, and then infers the spatial locations of additional cells by using their transcriptomic profiles generated by snRNA-seq.

We utilized LIBD ST data from 8 out of 12 sections of the DLPFC as the reference ST data for training the CeLEry model. We selected these 8 sections because they encompassed data of all six cortical layers and WM. Specifically, sections with IDs 151673, 151674, 151675, and 151676 were originated from the same brain sample. Sections 151507, 151508, 151509, and 151510 were originated from another brain. Since CelEry can utilize ST reference data profiled from either one or multiple sections, we conducted experiments to select the best reference for inferring the spatial location of cells in the ROS/MAP snRNA-seq dataset. We considered various strategies by using only a single section, multiple sections from the same brain, and multiple sections from both brains as the reference data.

To evaluate the accuracy of inferred spatial domains, we compared the inferred cell type proportions in cortical layers based on different reference ST data to the golden standard provided by the single-cell resolution CosMx ST data. We used Kullback–Leibler (KL) divergence (16) to measure the similarity between the matrix of cell type proportions in all six cortical layers and WM inferred by CelEry and the matrix based on the single-cell resolution CoxMx ST data. KL divergence, denoted as *n_kl_ (P||Q),* is a type of statistical distance, measuring how one probability distribution *P* is different from a second reference probability distribution *Q*.

Here, the probability distribution *P(x)* refers to the inferred cell type proportions for each cortical layer or WM summing up to 1, and the probability distribution *Q(x)* represents the cell type proportions for each cortical layer or WM of the single cell resolution CosMx ST data. Since *P(x)* and *Q(x)* represent columns in the comparing matrices, the KL divergence as calculated by the following equation (1) is appropriate for comparing the distance between two matrices with cell type proportions in all six cortical layers and WM in the columns.

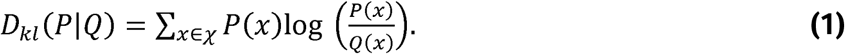

The reference ST data giving the cell type proportion matrix with the smallest KL divergence to the one based on CosMx ST data will be selected as the optimal ST reference data.

### CLS-CTS DGE analysis by GEMMA

With the inferred spatial locations for all cells in the ROS/MAP snRNA-seq data, we next conducted CLS-CTS DGE analyses for three quantitative AD-related phenotypes, β-amyloid, tangle density, and cognitive decline. For each gene of each sample, we quantified pseudo bulk read counts of each cell type in each cortical layer. Genes with average pseudo read counts less than five were excluded from the downstream analysis. Pseudo bulk read counts were normalized and converted to read counts per million base pairs (CPM). The log2 transformed CPM data were used to conduct CLS-CTS DGE analysis.

To control for potential false positive inflations (17), we utilized the LMM approach implemented in the GEMMA tool (8). Confounding covariates including study, age at death, sex, and post-mortem interval were adjusted for in the LMM. To test if gene *l* with expression levels *x*_j,k,l_ of cell type *k* in cortical layer *l* is differentially expressed with respect to a quantitative trait *v*_nx1_ (*H*_O_: *p*_j,k,l_ = 0), the following LMM is assumed:

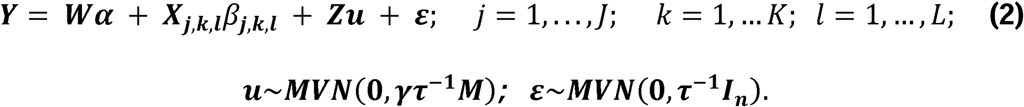

Here, *p*_j,k,l_ is the effect size of gene *j* in cell type *k* and cortical layer *l; W* is a *nxc* matrix of confounding covariates and an intercept term; *a* is a *cx t* vector of covariate effects; *Z* is a *nx n* loading matrix which is taken as an identify matrix (17); *e* is a *nx t* vector of independent errors following a normal distribution with mean 0 and variance *r*_-1_; ***u*** is a *nx t* vector denoting random effects of all *n* samples following a Multivariate Normal distribution with mean **0** and variance-covariance matrix *rr*_-1_*M; r* is the ratio of variance components between the random effects and error term; *M* is a *nx n* sample-sample correlation matrix derived from transcriptome-wide gene expression profiles; and *l*_n_ is a *nx n* identity matrix.

For comparison, we also conducted CTS only DGE analysis by using the same LMM model as in the above Equation (2), and pseudo bulk read counts obtained for each cell type per gene per sample. That is, for each gene *j*, we tested if its expression levels *x*_j,k_ in cell type *k* is differentially expressed with respect to *v*_nx1_ (*H*_O_: *p*_j,k_ = *0*).

### Gene Set Enrichment Analysis

Pre-ranked Gene set enrichment analysis (GSEA) as implemented in the *fgsea* (18) package in R makes it possible to identify pathways that contain many co-regulated genes from a priori defined gene sets (18). The pre-ranked GSEA method enhances the sensitivity and accuracy of pathway enrichment analyses in transcriptional datasets. For each phenotype, we conducted GSEA analyses by *fgsea* with top 500 significant genes from two pairs of cell type and cortical layer which have top CLS-CTS significant genes. We utilized multiple curated gene set data bases from the Human Molecular Signatures Database MSigDB (19) –– hallmark gene sets (H), curated gene sets (C2), ontology gene sets (C5) and cell type signature gene sets (C8). These hallmark gene sets summarize specific biological states or processes through coherent expression patterns, crafted by identifying overlaps across gene sets and retaining coordinately expressed genes. The curated gene sets are assembled from online pathway databases, biomedical literature, and inputs from domain experts, this collection includes gene sets categorized under chemical and genetic perturbations and canonical pathways. Ontology gene sets contain genes annotated by the same ontology term, with subcollections from Gene Ontology that cover biological processes, cellular components, and molecular functions, as well as from the human phenotype ontology. Cell type signature gene sets were derived from single-cell sequencing studies. These gene sets represent cluster marker genes for various cell types across multiple human tissues and are used to aid cell type assignment in complex datasets, such as those derived from organoid models.

## Results

### Accurate cortical layers were inferred for cells with snRNA-seq profiles

Inferred cortical layers were obtained for ∼1.5 million cells in the ROS/MAP snRNA-seq data of DLPFC, by using the CeLEry tool with different ST reference data (see Methods). The numbers and proportions of cells inferred to each cortical layer and WM were presented in **Supp Table 1.** The proportions of cells inferred to each cell type of each cortical layer and WM were plotted in the heatmaps in **Supp Fig 1** and bar plots in **Supp Fig 2.** We assessed the accuracy of predicted cell type proportions in cortical layers and WM by comparing their KL divergences to the cell type proportions in the CosMx ST data profiled from the same DLPFC tissue (see Methods). The minimum KL divergence was observed in the location inference results based on the reference ST data from sample 151673 (**Fig 2**).

**Fig 2:**
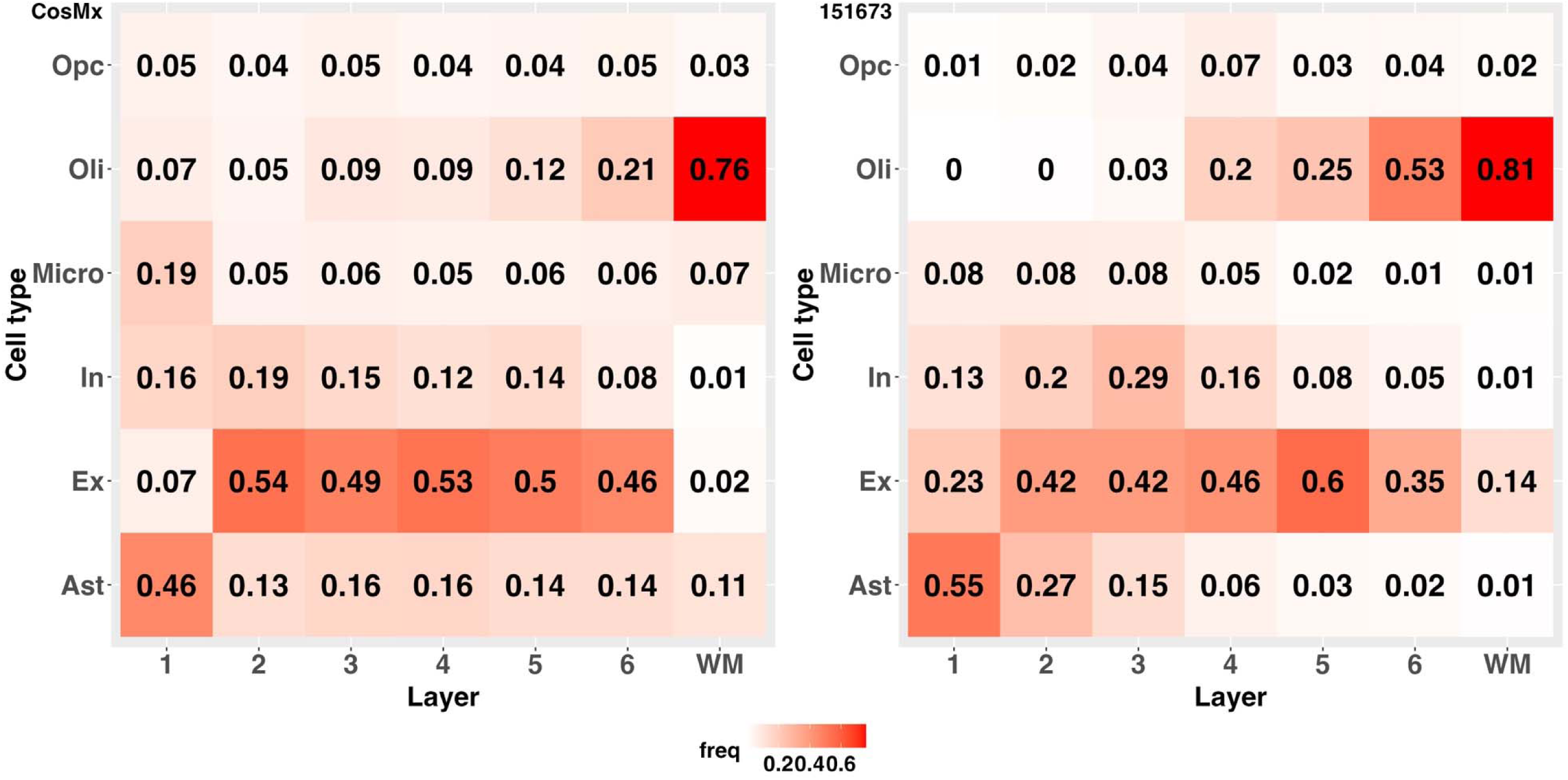
Cell type proportions in cortical layers and WM based on the single cell resolution CosMx ST data (left panel), and the inferred results for ROSMAP snRNA-seq data using LIBD reference sample 151673 (right panel).

From the cortical layers inferred based on reference sample 151673 (**Supp Fig 1**), we found cortical layer 3 contained most cells (26.08%) compared to other cortical layers. Also, from the right panel in **Fig 2**, we found that astrocyte is the major cell type in cortical layer 1 with proportion 55%; excitatory neuron was the major cell type in cortical layers 2-5; oligodendrocyte was the major cell type in cortical layers 6 (53%). These cell type proportions in the inferred cortical layers and WH are similar to the ones obtained from the single cell resolution CosMx ST data (left panel in **Fig2**), and consistent with the results shown by real application studies of DLPFC tissue in the CeLEry paper (7).

These inferred locations of cortical layers 1-6 were used in the follow-up CLS-CTS DGE analyses. Cells inferred to the WM layer were not considered in our CLS-CTS DGE analyses. Since cells inferred to the WM are primarily oligodendrocytes (81%), the number of cells is limited for studying other cell types in WM. Also, the results are expected to be similar as the ones obtained by CTS DGE analyses of oligodendrocytes.

### CLS-CTS DGE analyses identified cortical layer specific significant genes

We conducted the CTS-CTS DGE analyses for three quantitative AD-related phenotypes –– β-amyloid (n=433), tangle density (n=433), and cognitive decline (n=436). By utilizing the LMM, we observed that false positive rates were well calibrated compared to the results with severely inflated false positives obtained by standard linear regression modeling (**Supp Fig 3**) (17). We identified a total of 450 potential CLS-CTS significant genes with nominal p-values <10^-4^, including 258 for β-Amyloid, 122 for tangle density, and 127 for cognitive decline (**Fig 3A**). To show the improved power by incorporating inferred spatial location information in the DGE analyses, we compared to the results obtained by standard CTS DGE analyses with the same ROS/MAP snRNA-seq and phenotype data. The standard CTS DGE analyses identified 141 potential significant genes with p-values <10^-4^, including 53 for β-amyloid, 47 for tangle density, and 61 for cognitive decline. A shown in **Fig 3A**, 60.4% of these potential significant genes for β-amyloid, 51.1% for tangle density, and 55.7% for cognitive decline were also detected by CLS-CTS DGE analyses. Without incorporating spatial information, 226 significant genes in β-amyloid, 98 in tangle density, and 93 in cognitive decline were missed.

**Fig 3:**
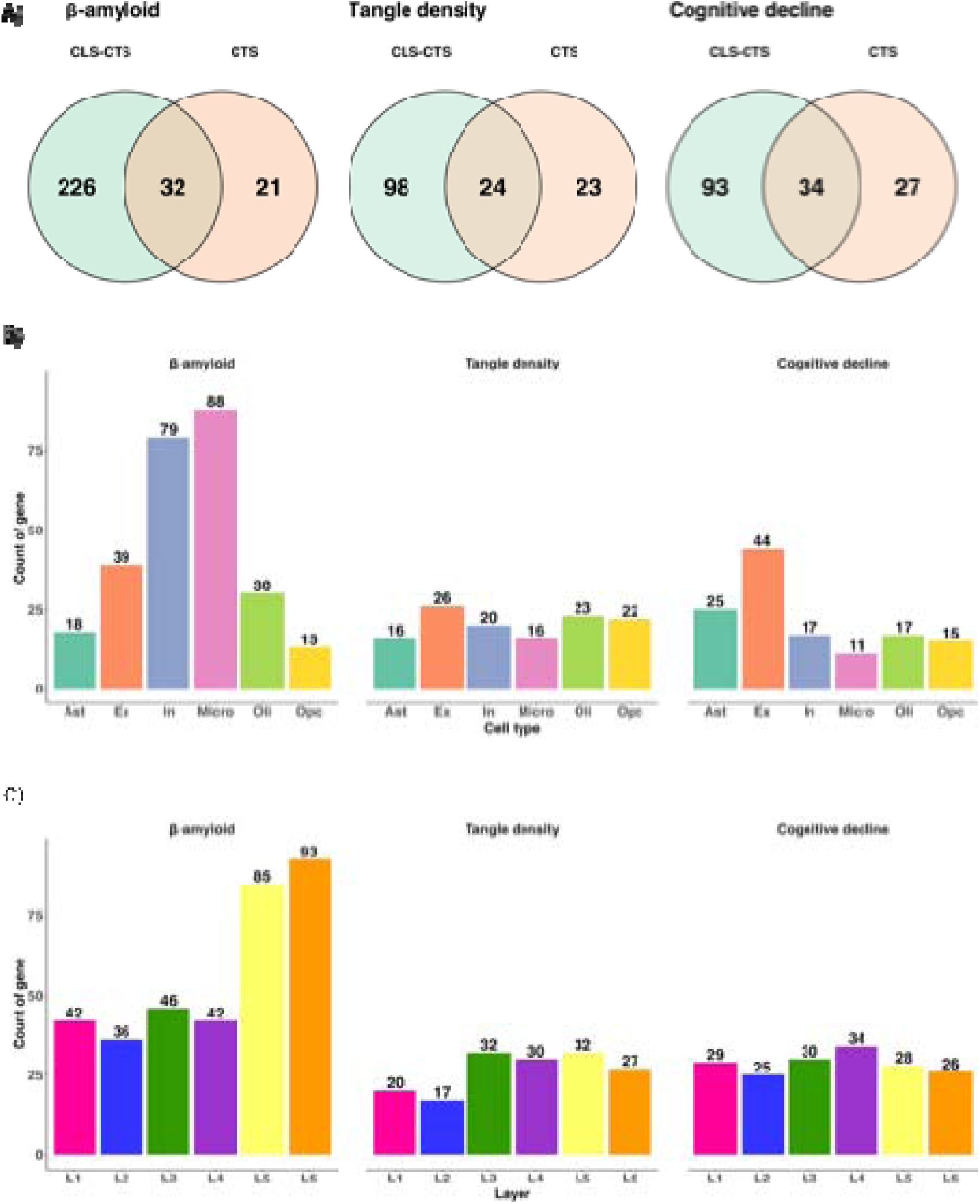
Results of CLS-CTS DGE analyses of P-amyloid, tangle density, and cognitive decline. A) Venn diagram of the numbers of CLS-CTS significant genes and CTS significant genes (with p-values < 10^-4^). Majority CLS-CTS significant genes were missed by standard CTS DGE analysis not considering spatial information. B) Bar chart representing the counts of CLS-CTS significant genes for each cell type. T The numbers of CLS-CTS significant genes of /3-amyloid are predominant in inhibitory neurons and Microglia cell types, while the ones of tangle density and cognitive decline are evenly distributed across all cell types. C) Bar chart representing the counts of CLS-CTS significant genes for cortical layers 1-6. The numbers of CLS-CTS significant genes of /3-amyloid are predominant in cortical layers 5 and 6, while the ones for tangle density and cognitive decline are evenly distributed across all layers.

We present the distributions of numbers of significant CLS-CTS genes of each AD-related phenotype across six cell types in **Fig 3B**, and across six cortical layers in **Fig 3C**. For β-amyloid, most significant CLS-CTS genes were observed in microglia (N=88) and inhibitory neurons (N=79), predominantly in cortical layers 5 and 6. For cognitive decline and tangle density, the numbers of significant CLS-CTS genes are evenly distributed across cell types and cortical layers with similar patterns. The largest number of significant genes for cognitive decline and tangle density is obtained in excitatory neurons and cortical layers 3-5.

### Gene Set Enrichment Analyses identified AD-related pathways

For each phenotype, we conducted four sets of GSEA analyses, each with selected top 500 significant genes in the CLS-CTS DGE results of the top two cortical layers and top two cell types that have the highest numbers of significant genes (**Table 1**). For β-amyloid, the top two cell types were identified as microglia and inhibitory neurons that contain most CLS-CTS significant genes. Then cortical layers 5 and 6 were identified as the top two cortical layers containing most CLS-CTS significant genes in microglia, and cortical layers 1 and 5 were identified for inhibitory neurons. For tangle density, cortical layers 5 and 6 in excitatory neurons and cortical layers 4 and 5 in oligodendrocytes were identified. Given the similarity in the distribution of significant CLS-CTS genes between cognitive decline and tangle density, we selected the same cell types of excitatory neurons (with most significant genes) in cortical layers 1 and 5 and oligodendrocytes (with third most significant genes) in cortical layers 5 and 6 for cognitive decline.

**Table 1:** Numbers of CLS-CTS significant genes in top two cell types of AD-related phenotypes. The top two numbers of significant genes are bold for each cell type and each phenotype.

| Phenotype | Cell type | L1 | L2 | L3 | L4 | L5 | L6 |
| --- | --- | --- | --- | --- | --- | --- | --- |
| β-amyloid | Microglia | 5 | 12 | 11 | 9 | **35** | **51** |
| β-amyloid | Inhibitory neurons | **18** | 16 | 15 | 15 | **18** | 0 |
| Tangle density | Excitatory neurons | 3 | 4 | 2 | 6 | **8** | **7** |
| Tangle density | Oligodenrocytes | 0 | 0 | 0 | **11** | **11** | 7 |
| Cognitive decline | Excitatory neurons | **15** | 11 | 5 | 6 | **12** | 8 |
| Cognitive decline | Oligodenrocytes | 0 | 0 | 2 | 2 | **5** | **10** |

We identified a total of 156 significant pathways (p-adj <0.05, adjusted for multiple testing) enriched with CLS-CTS significant genes of β-amyloid in cortical layer 6 of microglia. Among these, 12 pathways are directly related to AD, as shown in **Fig 4A**. Notably, ribosomal pathways such as the Ribosome from KEGG (p-adj= 1.64E-15) (20–22), Cytoplasmic ribosomal proteins from WikiPathways (p-adj=3.87E-13) (23), and Ribosome from GOCC (p-adj=4.73E-14) (24,25) were identified, underscoring the role of ribosome family proteins in AD pathogenesis (26).

**Fig 4:**
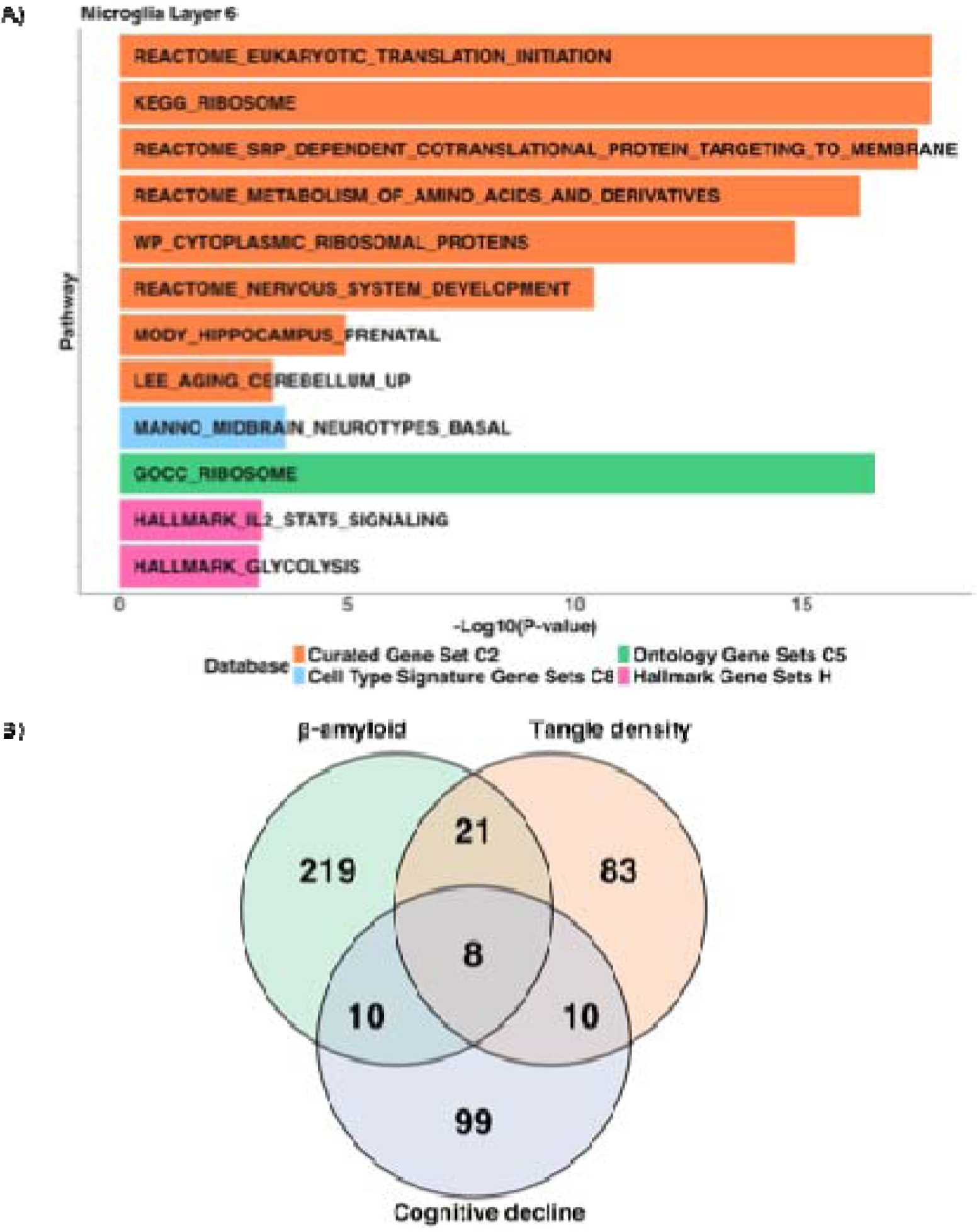
GSEA results and shared significant CLS-CTS genes across three analyzed AD-related phenotypes. A) AD-related significant pathways obtained by GSEA, with top 500 significant genes of /3-amyloid for Microglia in cortical Layer 6. B) Venn diagram showing the overlapped numbers of CLS-CTS significant genes (p-value < 10^-4^) across /3-amyloid, tangle density, and cognitive decline; 8 genes were found shared across all three phenotypes, 21 genes between /3-amyloid and tangle density, 10 genes between cognitive decline and tangle density, and 10 genes between cognitive decline and /3-amyloid.

Additional significant AD-related pathways identified in the curated gene sets C2 include: i) The Eukaryotic Translation Initiation pathway from REACTOME (p-adj=1.64E-15) (27) which underscores the role of the translational machinery, particularly the α subunit of eukaryotic initiation factor-2 (eIF2α), in AD pathogenesis (28). ii) Pathway SRP-dependent cotranslational protein targeting to membrane from REACTOME (p-adj=1.89E-14) focuses on SRP-dependent cotranslational protein targeting to the membrane. Proteins in this pathway have been associated with periodontal disease and AD, indicating a broader impact on health (29). iii) Pathway Metabolism of amino acids and derivatives from REACTOME (p-adj=1.64E-15) (30–32) covers the metabolism of amino acids and their derivatives, noting significant changes in the levels of enzymes involved in amino acid metabolism observed in the AD brain (33). iv) Pathway Nervous system development from REACTOME (p-adj=9.32E-9), which focuses on the developmental processes of the nervous system (34–36). v) Pathway of genes highly expressed in prenatal hippocampus from MODY (p-adj=1.12E-5), highlighting genes highly expressed in the prenatal hippocampus, reflecting the critical developmental molecular programs which is a cortical structure critical for learning and memory (37). vi) Pathway about upregulated in the cerebellum of aged adult mice compared to young adults from LEE (p-adj=0.038), is pathway highlighting how aging impacts cognitive and motor skills and is a risk factor for neurological disorders including AD and Parkinson’s disease (38).

One significant AD related pathway, Human embryonic midbrain neurotypes in basal plate from MANNO, was identified in the cell type signature gene sets C8. This pathway pertains to the molecular diversity of midbrain development across mouse, human, and stem cell studies (p-adj=4.58E-3) (39). Understanding the human embryonic ventral midbrain is crucial for research on Parkinson’s disease and may also intersect with AD mechanisms.

In addition, two significant AD pathways were identified in hallmark gene sets H. One Pathway was Genes encoding proteins involved in glycolysis and gluconeogenesis (p-adj=0.02) (19). Notably, reduced glucose metabolism is observed in patients at risk for AD, highlighting the relevance of this pathway (40). The second pathway Genes up-regulated by STAT5 in response to IL2 stimulation (p-adj=0.02) (19), where IL-2 plays a dual role by enhancing memory formation and controlling inflammation, making it a promising target for AD research (41).

In cognitive decline, total 5 significant pathways were identified, including one AD related Pathway that contains genes up-regulated by activation of the PI3K/AKT/mTOR pathway (p-adj=0.044) (19) in oligodendrocytes in cortical layer 6. This pathway is crucial in the development and progression of AD, offering potential therapeutic targets that could improve patient outcomes (42). For tangle density, only one significant Pathway Genes involved in the G2/M checkpoint (19), was identified in cortical layer 5 and another one Pathway Genes sets of stellate cell from human liver by Aizarani (43) in cortical layer 6 in excitatory neurons, which have no known association with AD.

As a comparison, we also conducted GSEA by using the top 500 significant genes obtained by CTS DGE analyses for each phenotype. Significant AD-related pathways are presented in **Supp Fig 4A.** These significant pathways are not overlapped with those identified based on CLS-CTS DGE analysis results, highlighting that CLS-CTS DGE provided additional insights for studying AD related phenotypes.

### Genes with CLS-CTS differential gene expression across multiple AD-related phenotypes

By comparing the CLS-CTS significant genes with differential expression for these three analyzed AD-related phenotypes, we found 8 significant genes shared by all three phenotypes, 21 genes shared by β-amyloid and tangle density, 10 genes shared by cognitive decline and tangle density, and 10 genes shared by β-amyloid and cognitive decline (**Fig 4B**; **Supp Figs 5-7**).

Although several significant genes such as *NPNT*, *CREB5*, *DPYD*, and *PTPRG* were also found significant for all three phenotypes by CTS DGE analyses (**Table 2**), we can detect in which cortical layers these genes have differential gene expression with respect to the phenotype of interest by incorporating the spatial location information. For instance, *CREB5*, pivotal in synaptic plasticity, was found significantly differentially expressed in astrocytes –– in cortical layer 3 for β-amyloid (p-value = 1.04E-07), cortical layers 1 and 2 for tangle density (p-value = 2.58E-07), and cortical layer 2 for cognitive decline (p-value = 3.80E-6). The *CREB* signaling is crucial for the conversion of short-term memory to long-term memory and is implicated in various cognitive and neuro DGE generative disorders (44). Importantly, 2 out of 8 genes were missed if only conducting CTS DGE analysis without considering the spatial location. For example, *KCNIP3* encoded by Calsenilin is a known AD risk factor (45), which was found significant in oligodendrocytes for all three AD-related phenotypes only in cortical layer 5, with p-value = 2.85E-05 for β-amyloid, p-value = 1.08E-06 for tangle density, and p-value = 2.45E-11 for cognitive decline.

**Table 2:**
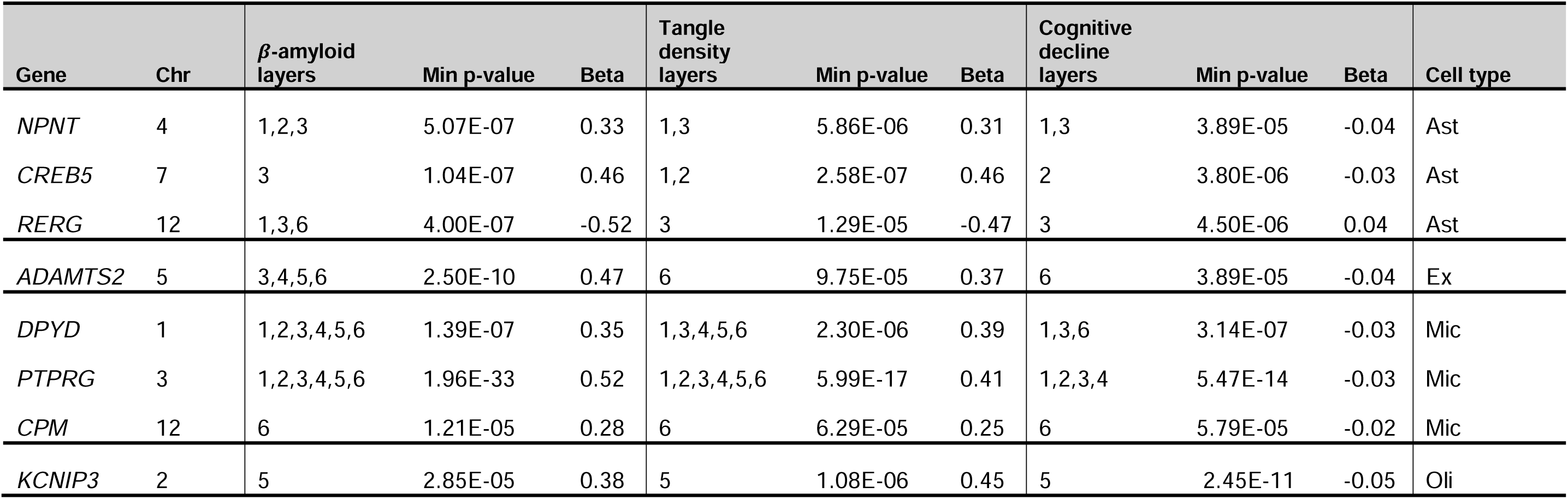
A total of 8 CLS-CTS significant genes (p-value < 10^-4^) shared across all three AD-related phenotypes. The columns ofa “phenotype-layers” indicate the layers where these genes have differential expression with respect to the phenotype. Columns of “Min p-value” and “Beta” indicate the minimum p-values across all layers, and the corresponding effect sizes. The “Cell type” column shows the cell type of which the genes have differential expression.

Similar results were also observed for shared significantly differentially expressed genes for β-amyloid and tangle density (**Table 3**). We found 15 out of 21 shared significant genes were missed by CTS DGE analyses. For example, the well-known AD-related gene *APOE* (46) was found significant in microglia only in cortical layers 5 and 6 for β-amyloid (p-value = 3.77E-06) and only in cortical layer 2 for tangle density (p-value = 1.22E-05). Also, *CTSD* that was found significant in microglia only in cortical layer 6 with p-value =1.44E-06 for β-amyloid and p-value = 8.28E-05 for tangle density, is known as a gene involved in amyloid precursor protein processing and a candidate gene for AD (47). The gene expressions of *APOE* and *CTSD* were not found differentially expressed by CTS DGE analyses.

**Table 3:** A total of 21 CLS-CTS significant genes (p-value < 10^-4^) shared across phenotypesβ-amyloid and tangle density. The columns of “phenotype-layers” indicate the layers where these genes have differential expression with respect to the phenotype. Columns of “Min p-value” and “Beta” indicate the minimum p-values across all layers, and the corresponding effect sizes. The “Cell type” column shows the cell type of which the genes have differential expression. Two cell types in “Cell type” column indicates the gene show significant across different cell types for different phenotypes, the first one is for β-amyloid, the second one is for tangle density.

| Gene name | Chr | β-amyloid layer | Min p-value | Beta | Tangle density layer | Min p-value | Beta | Cell type |
| --- | --- | --- | --- | --- | --- | --- | --- | --- |
| *ATP8A* | 4 | 1 | 4.70E-05 | -0.87 | 2 | 1.39E-05 | -1.10 | Ast |
| *AC016831.5* | lnc-RNA | 1,2,3,3,5 | 1.10E-09 | 0.91 | 3,5 | 4.02E-05 | 0.83 | Ex, In |
| *AC093607.1* | lnc-RNA | 3,4,5,6 | 3.24E-07 | -0.31 | 4 | 8.82E-05 | -0.28 | Ex |
| *STRADB* | 2 | 1,2,4,5 | 8.43E-08 | -0.59 | 3 | 2.39E-05 | -0.49 | In, Opc |
| *AC211433.1* | lnc-RNA | 1,2 | 7.30E-06 | -0.29 | 3 | 3.42E-05 | -0.31 | In, Opc |
| *MYCBP2-AS1* | 13 | 1 | 4.00E-06 | -0.33 | 3 | 9.87E-05 | -0.33 | In, Opc |
| *HMGN1* | 21 | 2 | 9.69E-05 | -0.49 | 3 | 3.18E-05 | -0.47 | In, Opc |
| *TENM3-AS1* | 4 | 4,5 | 2.19E-06 | -0.46 | 6 | 9.89E-05 | -0.29 | In |
| *HMGN2* | 1 | 6 | 5.48E-06 | -0.46 | 2 | 8.93E-05 | -0.75 | In, Ex |
| *FOXP1* | 3 | 2,3,5,6 | 1.79E-07 | 0.65 | 1,2,3,4,5 | 9.66E-11 | 0.70 | Mic |
| *IL15* | 4 | 2,4 | 9.42E-06 | 0.36 | 2,4 | 2.11E-06 | 0.39 | Mic |
| *ZNF618* | 9 | 2 | 1.77E-05 | 0.49 | 4 | 3.84E-05 | 0.45 | Mic |
| *CADM1* | 11 | 3 | 3.56E-05 | 0.43 | 4 | 4.29E-06 | 0.46 | Mic |
| *APOE* | 19 | 5,6 | 3.77E-06 | 0.25 | 2 | 1.22E-05 | 0.47 | Mic |
| *CTSD* | 11 | 6 | 1.44E-06 | 0.35 | 6 | 8.28E-05 | 0.42 | Mic |
| *SLC6A9* | 1 | 4,5 | 2.63E-05 | 0.42 | 5 | 3.12E-08 | 0.54 | Oli |
| *AC093865.1* | lnc-RNA | 4 | 9.93E-05 | 0.26 | 4 | 7.21E-05 | 0.27 | Oli |
| *SLC38A2* | 12 | 5 | 1.75E-05 | 0.33 | 5 | 6.26E-05 | 0.32 | Oli |
| *METTL7A* | 12 | 5 | 5.29E-05 | 0.37 | 6 | 6.54E-05 | 0.34 | Oli, Opc |
| *AC009063.2* | lnc-RNA | 5 | 2.01E-05 | 0.30 | 4 | 3.17E-06 | 0.35 | Oli |
| *MID1IP1* | X | 6 | 2.93E-06 | 0.38 | 5,6 | 2.88E-07 | 0.43 | Oli |

Among the shared significantly differentially expressed genes for β-amyloid and cognitive decline (**Table 4**), We found 5 out of 10 shared significant genes were missed by CTS DGE analyses. For example, *ALCAM* that was found significant in microglia only in cortical layer 5 for β-amyloid (p-value = 9.33E-06) and only in cortical layers 3 and 6 for cognitive decline (p-value = 1.69E-06), is indicative of increased AD risk when elevated in plasma (48).

**Table 4:** A total of 10 CLS-CTS significant genes (p-value < 10^-4^) shared across the phenotypes:β-amyloid and cognitive decline. The columns of “phenotype-layers” indicate the layers where these genes have differential expression with respect to the phenotype. Columns of “Min p-value” and “Beta” indicate the minimum p-values across all layers, and the corresponding effect sizes. The “Cell type” column shows the cell type of which the genes have differential expression.

| Gene name | Chr | β-amyloid layer | Min p-value | Beta | Cognitive decline layer | Min p-value | Beta | Cell type |
| --- | --- | --- | --- | --- | --- | --- | --- | --- |
| *AGBL1* | 15 | 2,4 | 3.58E-06 | -0.28 | 5 | 1.85E-06 | 0.03 | Ex |
| *AL122014.1* | lnc-RNA | 2,3 | 4.61E-07 | -0.34 | 1,2,3 | 4.70E-09 | 0.04 | Ex |
| *AC015943.1* | lnc-RNA | 6 | 1.27E-07 | -0.33 | 6 | 9.16E-06 | 0.03 | Ex |
| *AC034268.2* | lnc-RNA | 6 | 9.05E-05 | -0.31 | 1,6 | 7.91E-07 | 0.04 | Ex |
| *HES4* | 1 | 5 | 2.58E-06 | -0.39 | 4 | 7.40E-09 | 0.04 | In |
| *SPRY4-AS1* | 5 | 5,6 | 4.53E-06 | -0.23 | 5,6 | 1.50E-06 | 0.02 | In |
| *ALCAM* | 3 | 5 | 9.33E-06 | 0.26 | 3,6 | 1.69E-06 | -0.03 | Mic |
| *PRKCH* | 14 | 6 | 1.22E-05 | 0.32 | 4 | 8.80E-05 | -0.04 | Mic |
| *HS3ST4* | 16 | 5 | 4.66E-05 | 0.20 | 4 | 7.07E-05 | -0.03 | Mic |
| *P3H2* | 3 | 6 | 1.48E-05 | 0.36 | 6 | 1.52E-05 | -0.03 | Oli |

Among the shared significantly differentially expressed genes for tangle density and cognitive decline (**Table 5**), We found 5 out of 10 shared significant genes were missed by CTS DGE analyses. For example, *PFKP* that was found significant in astrocytes only in cortical layer 6 for tangle density (p-value = 1.28E-07) and cognitive decline (p-value = 1.20E-05), has been associated with AD-specific impairments in glucose metabolism (49). *AMGO* has been implicated in neurodevelopmental disorders, including autism spectrum disorders (50), which was found significant in oligodendrocyte precursor cells only in cortical layer 5 for tangle density (p-value = 3.21E-05) and cortical layer 4 and 5 for cognitive decline (p-value = 4.79E-05).

**Table 5:** A total of 10 CLS-CTS significant genes (p-value < 10^-4^) shared across the phenotypes: tangle density, and cognitive decline. The columns of “phenotype-layers” indicate the layers where these genes have differential expression with respect to the phenotype. Columns of “Min p-value” and “Beta” indicate the minimum p-values across all layers, and the corresponding effect sizes. The “Cell type” column shows the cell type of which the genes have differential expression. Two cell types in “Cell type” column indicates the gene show significant across different cell types for different phenotypes, the first one is for tangle density, the second one is for cognitive decline.

| Gene name | Chr | Tangle density layer | Min p-value | Beta | Cognitive decline layer | Min p-value | Beta | Cell type |
| --- | --- | --- | --- | --- | --- | --- | --- | --- |
| *OXR1* | 8 | 1 | 9.28E-05 | 0.84 | 3 | 6.20E-05 | -0.04 | Ast, Mic |
| *PFKP* | 10 | 6 | 1.28E-07 | 0.45 | 6 | 1.20E-05 | -0.03 | Ast |
| *AC021733.4* | lnc-RNA | 5 | 5.97E-06 | -0.73 | 3 | 9.13E-06 | 0.03 | Ex, In |
| *RPH3A* | 12 | 6 | 3.35E-05 | -0.81 | 6 | 2.45E-06 | 0.09 | Ex |
| *PLA2G4C* | 19 | 1 | 1.27E-06 | -0.52 | 1,4 | 2.21E-06 | 0.05 | In |
| *PLD5* | 1 | 4,5 | 2.85E-06 | 0.50 | 4,5 | 1.60E-05 | -0.04 | Oli |
| *ASPA* | 17 | 5,6 | 3.68E-05 | -0.54 | 3,6 | 8.00E-07 | 0.04 | Oli |
| *VCAN* | 5 | 3,4 | 3.53E-06 | 0.73 | 2,3 | 1.28E-05 | -0.03 | Opc, Ast |
| *AGMO* | 7 | 5 | 3.21E-05 | 0.33 | 4,5 | 4.79E-07 | -0.03 | Opc |
| *AC007106.2* | lnc-RNA | 3,4,5,6 | 5.11E-07 | 0.40 | 4 | 5.62E-05 | -0.03 | Opc |

## Discussion

In this study, we designed a robust workflow that could integrate spatial ST data with snRNA-seq data to conduct CLS-CTS DGE analyses by LMM (8), for the goal of identifying spatially-informed differentially expressed genes for the phenotype of interest. We integrated the LIBD ST reference data (6) with ROS/MAP snRNA-seq data with large sample size (n=436) of DLPFC (3,4) to conduct spatially-informed DGE analyses for three AD-related phenotypes –– β-amyloid, tangle density, and cognitive decline. As a result, we identified 450 CLS-CTS significant genes, including 258 for β-amyloid, 122 for tangle density, and 127 for cognitive decline. Importantly, traditional CTS DGE analyses without spatial layer information failed to detect most of these differentially expressed genes, including 226 for β-amyloid, 98 for tangle density, and 93 for cognitive decline. Notably, *APOE*, a well-known AD-related gene, was only found significant in microglia in cortical layer 5 for β-amyloid and cortical layer 2 for tangle density, but not by CTS DGE analyses.

By GSEA analyses with top 500 significant CLS-CTS genes for top two cortical layers of top two cell types that have most significant genes for each phenotype, we found 156 significant pathways for β-amyloid, 2 for tangle density, and 5 for cognitive decline. Among those significant pathways for β-amyloid, we found 12 AD-related pathways by using the top significant CLS-CTS genes identified in microglia in cortical layer 6, illustrating potential molecular mechanisms through which microglial cells may influence β-amyloid. These findings are crucial for they illuminate specific cellular interactions and molecular pathways that could be targeted for therapeutic interventions for AD. Importantly, all of these 12 AD-related pathways were missed by GSEA analyses using the CTS DGE analyses results for β-amyloid.

By comparing the CLS-CTS significant genes across those three AD-related phenotypes, we identified 8 significant genes shared across all three AD-related phenotypes, 10 shared between β-amyloid and cognitive decline, 10 shared between tangle density and cognitive decline, 21 shared between β-amyloid and tangle density. Several AD-related CLS-CTS significant genes that have differential expression in multiple AD related phenotypes were missed by CTS DGE analysis alone. For example, gene *KCNIP3*, a recognized AD risk factor, consistently emerged as significant in oligodendrocytes across all phenotypes within cortical layer 5, underscoring its impact down to specific cortical layers. The most well-known AD-related gene *APOE,* significant in microglia in cortical layers 5 and 6 for β-amyloid and cortical layer 2 for tangle density. Our findings show that it is important to study how genes spatially interact with AD phenotypes by considering the spatial information, which would provide deeper insights into the roles of significant genes in disease progression.

These spatially-informed DGE results of three AD-related phenotypes enhance our understanding of the significant genes, by providing information about their differential expression in specific cell types and in specific cortical layers. Additional biological mechanisms of AD are informed, by knowing the specific cell types and specific cortical layers where genes have differential gene expression with respect to β-amyloid, tangle density, and cognitive decline, especially for known AD risk genes. We anticipate that this approach will uncover novel insights into the spatial distribution of gene expression changes associated with AD, offering a more detailed view of the cellular interactions and molecular pathways involved in AD progression compared to traditional CTS DGE analysis. This comprehensive study holds the potential to identify new therapeutic targets and contribute to the development of more effective interventions for AD.

### Conclusions

By integrating ST data and snRNA-seq data to conduct spatially-informed DGE analyses of AD-related phenotypes, we identified important CLS-CTS significant genes with differential expression in specific cell types and cortical layers for AD-related phenotypes. Important AD-related pathways were found enriched with our identified CLS-CTS significant genes, and several known AD risk genes were found as CLS-CTS significant genes for multiple AD-related phenotypes. This integration approach addressed the current challenges of conducting spatially-informed DGE analysis with ST data of limited sample sizes, or snRNA-seq data of larger sample sizes but lacking spatially information. Enhanced power is achieved for spatially informed DGE analyses by our approach, as shown by our real studies of AD-related phenotypes. This approach not only promises broader applications in both clinical and research settings, but also paves the way for developing more personalized and effective therapeutic strategies for AD.

## List of abbreviations

ST: Spatial transcriptomics
AD: Alzheimer’s disease
DGE: differential gene expression
snRNA-seq: single-nucleus RNA sequencing
DLPFC: dorsolateral prefrontal cortex
CLS: cortical-layer specific
CTS: cell-type specific
LIBD: Lieber Institute for Brain Development
ROS/MAP: the Religious Order Study (ROS) and the Rush Memory and Aging Project
Ast: astrocytes
Ex: excitatory neurons
In: inhibitory neurons
Mic: microglia
Oli: oligodendrocytes
Opc: oligodendrocyte precursor cells
AD-NC: Alzheimer’s disease neuropathologic changes
LMM: Linear mixed model Kullback–Leibler divergence
KL: divergence
GSEA: Gene Set Enrichment Analysis

## Declarations

### Ethics approval and consent to participate

All ROSMAP participants enrolled without known dementia and agreed to detailed clinical evaluation and brain donation at death [https://pubmed.ncbi.nlm.nih.gov/29865057/]. Both studies were approved by an Institutional Review Board of Rush University Medical Center (ROS IRB# L91020181, MAP IRB# L86121802). Both studies were conducted according to the principles expressed in the Declaration of Helsinki. Each participant signed an informed consent, Anatomic Gift Act, and an RADC Repository consent (IRB# L99032481) allowing her data and biospecimens to be repurposed. ROS/MAP transcriptomics data were shared with a data use agreement.

### Consent for publication

Not applicable.

### Authors’ contributions

S.T. conducted data analysis and drafted manuscript. S.L. annotated the cortical layer for CosMx ST data. A.S.B., D.A.B., P.L.D collected the ROS/MAP transcriptomic data, participated in data interpretation, and revised manuscript. J.H. and J.Y. conceived this study, supervised data analysis, data interpretation, and revised manuscript.

### Data availability

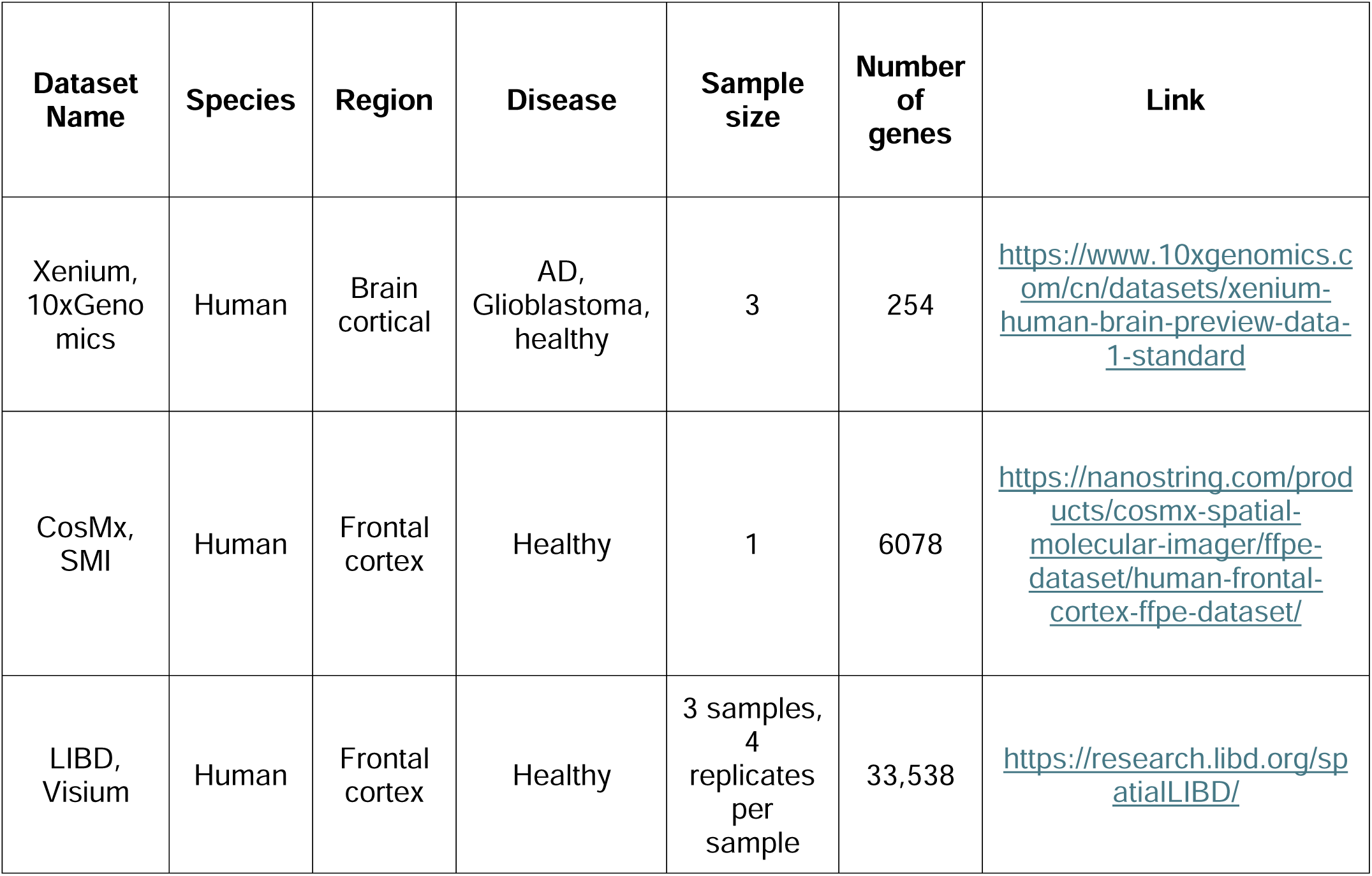

ROSMAP snRNA-Seq data request: https://www.synapse.org/Synapse:syn23650894

CelEry tool: https://github.com/QihuangZhang/CeLEry

Analysis scripts for this study are shared through the github directory: https://github.com/tangjiji199645/CLS-CTS-DGE

## Reference

1. Cao J, Li C, Cui Z, Deng S, Lei T, Liu W, et al. Spatial Transcriptomics: A Powerful Tool in Disease Understanding and Drug Discovery. Theranostics. 2024 May 11;14(7):2946–68.

2. Jaume G, Doucet P, Song AH, Lu MY, Almagro-Pérez C, Wagner SJ, et al. HEST-1k: A Dataset for Spatial Transcriptomics and Histology Image Analysis [Internet]. arXiv; 2024 [cited 2024 Sep 30]. Available from: http://arxiv.org/abs/2406.16192

3. Mathys H, Peng Z, Boix CA, Victor MB, Leary N, Babu S, et al. Single-cell atlas reveals correlates of high cognitive function, dementia, and resilience to Alzheimer’s disease pathology. Cell. 2023 Sep 28;186(20):4365–4385.e27.

4. Bennett DA, Buchman AS, Boyle PA, Barnes LL, Wilson RS, Schneider JA. Religious Orders Study and Rush Memory and Aging Project. J Alzheimers Dis JAD. 2018;64(Suppl 1):S161–89.

5. Piwecka M, Rajewsky N, Rybak-Wolf A. Single-cell and spatial transcriptomics: deciphering brain complexity in health and disease. Nat Rev Neurol. 2023 Jun;19(6):346–62.

6. Maynard KR, Collado-Torres L, Weber LM, Uytingco C, Barry BK, Williams SR, et al. Transcriptome-scale spatial gene expression in the human dorsolateral prefrontal cortex. Nat Neurosci. 2021 Mar;24(3):425–36.

7. Zhang Q, Jiang S, Schroeder A, Hu J, Li K, Zhang B, et al. Leveraging spatial transcriptomics data to recover cell locations in single-cell RNA-seq with CeLEry. Nat Commun. 2023 Jul 8;14(1):4050.

8. Zhou X, Stephens M. Genome-wide efficient mixed-model analysis for association studies. Nat Genet. 2012 Jun 17;44(7):821–4.

9. Pardo B, Spangler A, Weber LM, Page SC, Hicks SC, Jaffe AE, et al. spatialLIBD: an R/Bioconductor package to visualize spatially-resolved transcriptomics data. BMC Genomics. 2022 Jun 10;23(1):434.

10. V.S. Ramachandran, editor. Encyclopedia of the Human Brain [Internet]. Elsevier Science; 2002 [cited 2024 Aug 30]. Available from: http://www.sciencedirect.com:5070/referencework/9780122272103/encyclopedia-of-the-human-brain

11. Hu J, Li X, Coleman K, Schroeder A, Ma N, Irwin DJ, et al. SpaGCN: Integrating gene expression, spatial location and histology to identify spatial domains and spatially variable genes by graph convolutional network. Nat Methods. 2021 Nov;18(11):1342–51.

12. De Jager PL, Shulman JM, Chibnik LB, Keenan BT, Raj T, Wilson RS, et al. A genome-wide scan for common variants affecting the rate of age-related cognitive decline. Neurobiol Aging. 2012 May;33(5):1017.e1–15.

13. Boyle PA, Wang T, Yu L, Wilson RS, Dawe R, Arfanakis K, et al. To what degree is late life cognitive decline driven by age-related neuropathologies? Brain J Neurol. 2021 Aug 17;144(7):2166–75.

14. Bennett DA, Wilson RS, Schneider JA, Evans DA, Aggarwal NT, Arnold SE, et al. Apolipoprotein E epsilon4 allele, AD pathology, and the clinical expression of Alzheimer’s disease. Neurology. 2003 Jan 28;60(2):246–52.

15. Bennett DA, Schneider JA, Wilson RS, Bienias JL, Arnold SE. Neurofibrillary tangles mediate the association of amyloid load with clinical Alzheimer disease and level of cognitive function. Arch Neurol. 2004 Mar;61(3):378–84.

16. Csiszar I. $I$-Divergence Geometry of Probability Distributions and Minimization Problems. Ann Probab. 1975 Feb;3(1):146–58.

17. Tang S, Buchman AS, Wang Y, Avey D, Xu J, Tasaki S, et al. Differential gene expression analysis based on linear mixed model corrects false positive inflation for studying quantitative traits. Sci Rep. 2023 Oct 3;13(1):16570.

18. Korotkevich G, Sukhov V, Sergushichev A. Fast gene set enrichment analysis [Internet]. bioRxiv; 2019 [cited 2024 Jun 26]. p. 060012. Available from: https://www.biorxiv.org/content/10.1101/060012v2

19. Liberzon A, Birger C, Thorvaldsdóttir H, Ghandi M, Mesirov JP, Tamayo P. The Molecular Signatures Database (MSigDB) hallmark gene set collection. Cell Syst. 2015 Dec 23;1(6):417–25.

20. Kanehisa M, Goto S. KEGG: kyoto encyclopedia of genes and genomes. Nucleic Acids Res. 2000 Jan 1;28(1):27–30.

21. Kanehisa M, Furumichi M, Sato Y, Kawashima M, Ishiguro-Watanabe M. KEGG for taxonomy-based analysis of pathways and genomes. Nucleic Acids Res. 2023 Jan 6;51(D1):D587–92.

22. Kanehisa M. Toward understanding the origin and evolution of cellular organisms. Protein Sci Publ Protein Soc. 2019 Nov;28(11):1947–51.

23. Agrawal A, Balcı H, Hanspers K, Coort SL, Martens M, Slenter DN, et al. WikiPathways 2024: next generation pathway database. Nucleic Acids Res. 2024 Jan 5;52(D1):D679–89.

24. Ashburner M, Ball CA, Blake JA, Botstein D, Butler H, Cherry JM, et al. Gene Ontology: tool for the unification of biology. Nat Genet. 2000 May;25(1):25–9.

25. The Gene Ontology Consortium, Aleksander SA, Balhoff J, Carbon S, Cherry JM, Drabkin HJ, et al. The Gene Ontology knowledgebase in 2023. Genetics. 2023 May 2;224(1):iyad031.

26. Wang Y, Zhan D, Wang L. Ribosomal proteins are blood biomarkers and associated with CD4+ T cell activation in Alzheimer’s disease: a study based on machine learning strategies and scRNA-Seq data validation. Am J Transl Res. 2023 Apr 15;15(4):2498–514.

27. Merrick WC. Eukaryotic Protein Synthesis: Still a Mystery. J Biol Chem. 2010 Jul 9;285(28):21197–201.

28. Ohno M. Roles of eIF2α kinases in the pathogenesis of Alzheimer’s disease. Front Mol Neurosci. 2014 Apr 16;7:22.

29. Patel S, Howard D, Chowdhury N, Derieux C, Wellslager B, Yilmaz Ö, et al. Characterization of Human Genes Modulated by Porphyromonas gingivalis Highlights the Ribosome, Hypothalamus, and Cholinergic Neurons. Front Immunol. 2021 Jun 14;12:646259.

30. Häussinger D. Liver glutamine metabolism. JPEN J Parenter Enteral Nutr. 1990;14(4 Suppl):56S–62S.

31. Felig P. Amino acid metabolism in man. Annu Rev Biochem. 1975;44:933–55.

32. Owen OE, Reichard GA, Patel MS, Boden G. Energy metabolism in feasting and fasting. Adv Exp Med Biol. 1979;111:169–88.

33. Griffin JWD, Bradshaw PC. Amino Acid Catabolism in Alzheimer’s Disease Brain: Friend or Foe? Oxid Med Cell Longev. 2017;2017(1):5472792.

34. Kriegstein A, Alvarez-Buylla A. The glial nature of embryonic and adult neural stem cells. Annu Rev Neurosci. 2009;32:149–84.

35. Götz M, Huttner WB. The cell biology of neurogenesis. Nat Rev Mol Cell Biol. 2005 Oct;6(10):777–88.

36. Yao B, Christian KM, He C, Jin P, Ming GL, Song H. Epigenetic mechanisms in neurogenesis. Nat Rev Neurosci. 2016 Sep;17(9):537–49.

37. Mody M, Cao Y, Cui Z, Tay KY, Shyong A, Shimizu E, et al. Genome-wide gene expression profiles of the developing mouse hippocampus. Proc Natl Acad Sci U S A. 2001 Jul 17;98(15):8862–7.

38. Lee CK, Weindruch R, Prolla TA. Gene-expression profile of the ageing brain in mice. Nat Genet. 2000 Jul;25(3):294–7.

39. La Manno G, Gyllborg D, Codeluppi S, Nishimura K, Salto C, Zeisel A, et al. Molecular Diversity of Midbrain Development in Mouse, Human, and Stem Cells. Cell. 2016 Oct 6;167(2):566–580.e19.

40. Duran-Aniotz C, Hetz C. Glucose Metabolism: A Sweet Relief of Alzheimer’s Disease. Curr Biol. 2016 Sep 12;26(17):R806–9.

41. Alves S, Churlaud G, Audrain M, Michaelsen-Preusse K, Fol R, Souchet B, et al. Interleukin-2 improves amyloid pathology, synaptic failure and memory in Alzheimer’s disease mice. Brain. 2017 Mar 1;140(3):826–42.

42. Limantoro J, de Liyis BG, Sutedja JC. Akt signaling pathway: a potential therapy for Alzheimer’s disease through glycogen synthase kinase 3 beta inhibition. Egypt J Neurol Psychiatry Neurosurg. 2023 Nov 7;59(1):147.

43. Aizarani N, Saviano A, Sagar null, Mailly L, Durand S, Herman JS, et al. A human liver cell atlas reveals heterogeneity and epithelial progenitors. Nature. 2019 Aug;572(7768):199–204.

44. Saura CA, Valero J. The role of CREB signaling in Alzheimer’s disease and other cognitive disorders. Rev Neurosci. 2011;22(2):153–69.

45. Jo DG, Lee JY, Hong YM, Song S, Mook-Jung I, Koh JY, et al. Induction of pro-apoptotic calsenilin/DREAM/KChIP3 in Alzheimer’s disease and cultured neurons after amyloid-beta exposure. J Neurochem. 2004 Feb;88(3):604–11.

46. Kim J, Basak JM, Holtzman DM. The Role of Apolipoprotein E in Alzheimer’s Disease. Neuron. 2009 Aug 13;63(3):287–303.

47. Schuur M, Ikram MA, van Swieten JC, Isaacs A, Vergeer-Drop JM, Hofman A, et al. Cathepsin D gene and the risk of Alzheimer’s disease: a population-based study and meta-analysis. Neurobiol Aging. 2011 Sep;32(9):1607–14.

48. Chen J, Dai AX, Tang HL, Lu CH, Liu HX, Hou T, et al. Increase of ALCAM and VCAM-1 in the plasma predicts the Alzheimer’s disease. Front Immunol. 2022;13:1097409.

49. Saito ER, Miller JB, Harari O, Cruchaga C, Mihindukulasuriya KA, Kauwe JSK, et al. Alzheimer’s disease alters oligodendrocytic glycolytic and ketolytic gene expression. Alzheimers Dement. 2021;17(9):1474–86.

50. Sailer S, Keller MA, Werner ER, Watschinger K. The Emerging Physiological Role of AGMO 10 Years after Its Gene Identification. Life. 2021 Jan 26;11(2):88.

