## Supplemental for "Integrating spatial transcriptomics and snRNA-seq data enhances differential gene expression analysis results of AD-related phenotypes"

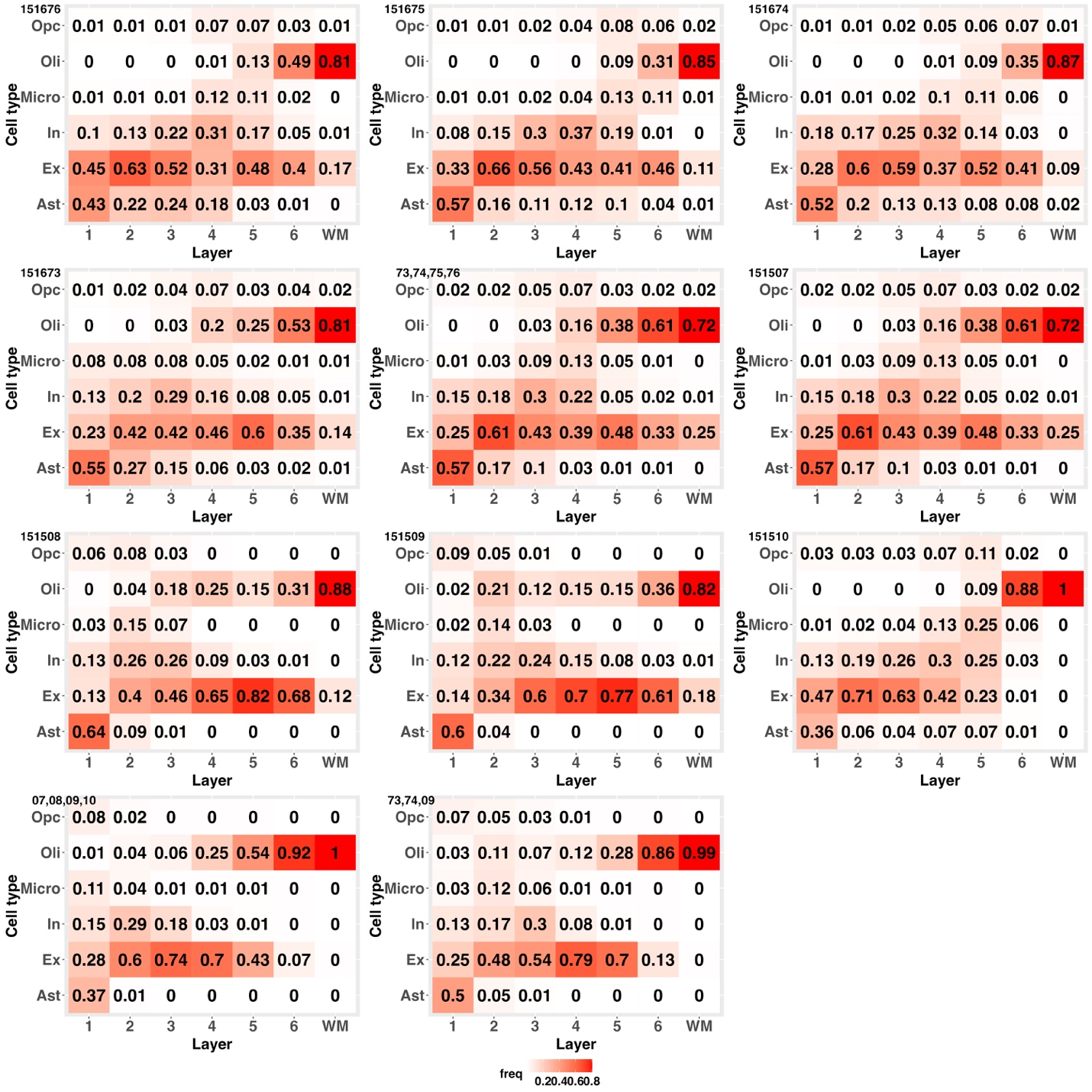

**Supp Fig 1.** **Cell type proportions of the ROS/MAP snRNA-seq data in each cortical layer and WM that were inferred by CeLEry based on different reference ST data.** Reference ST samples 151673, 151674, 151675, 151676 are sections from one brain, while samples 151507, 151508, 151509, 151510 are sections from another brain. Multiple section samples from the same brain were combined as one reference “sample 73,74,75,76” and “sample 07,08,09,10”. Multiple section samples (151673, 151674, and 151509) from different brains were combined as one reference “sample 73,74,09”.

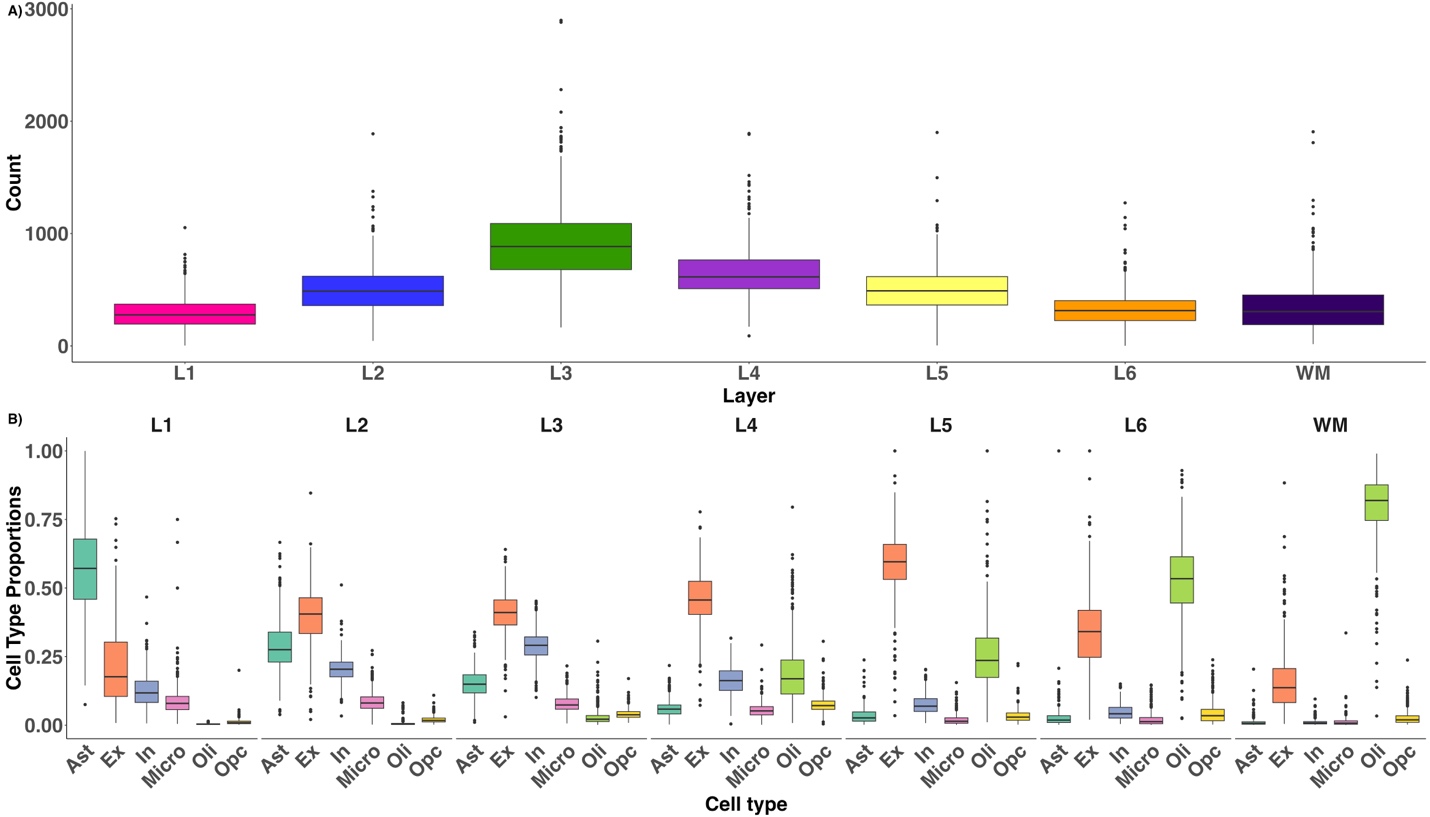

**Supp Fig 2. Distributions of annotated cells in the ROS/MAP sn-RNAseq data of 436 samples in six cortical layers and WM.** Spatial locations were inferred based on the reference ST sample 151673. (A). Distribution of cell counts across all six cortical layers and WM. (B). Cell type proportions for each layer and WM.

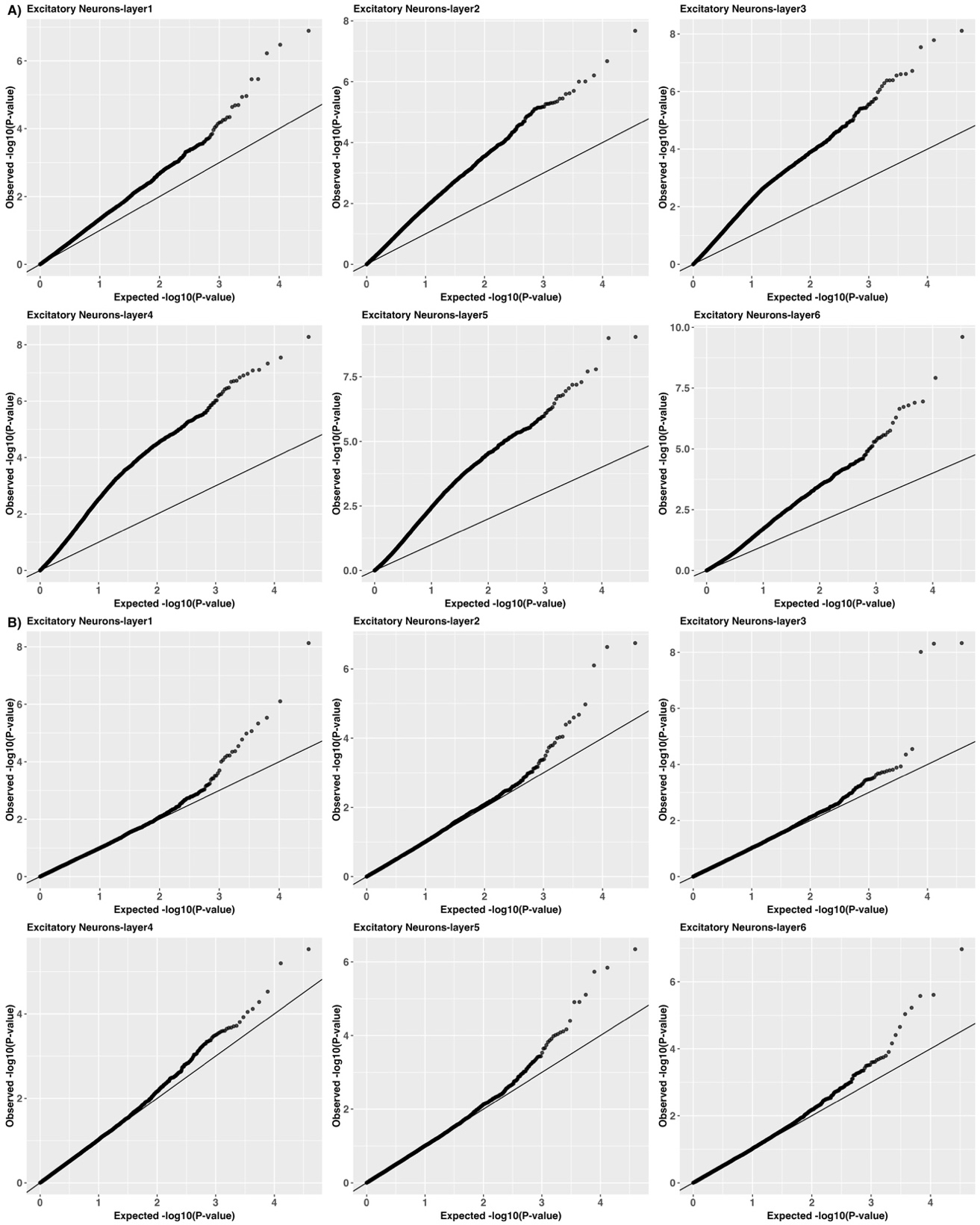

**Supp Fig3**. **Quantile-Quantile plots of log10(p-values) by CLS-CTS DGE analyses of** $\boldsymbol{\beta}$**-amyloid, for excitatory neurons in cortical layer 6 by standard linear regression model and LMM.** Strong inflation of false positive rates was observed in the results obtained by standard linear regression model (A), which were shown well calibrated in the results obtained by LMM (B).

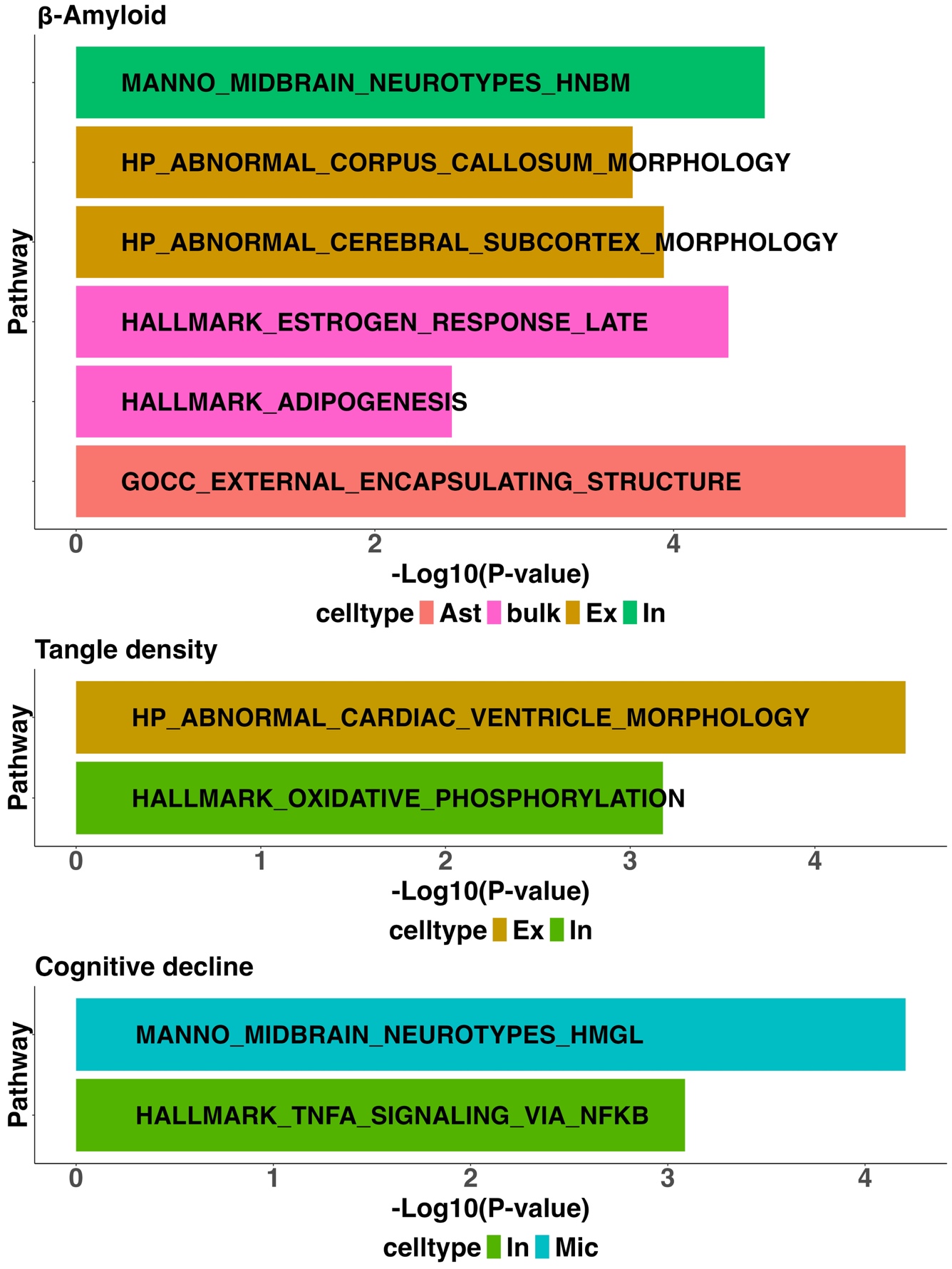

**Supp Fig 4. Significant AD related pathways identified by GSEA with CTS DGE results.** For each phenotype of β-amyloid, tangle density, and cognitive decline, top 500 significant genes selected from the CTS DGE results of each cell type were used for GSEA.

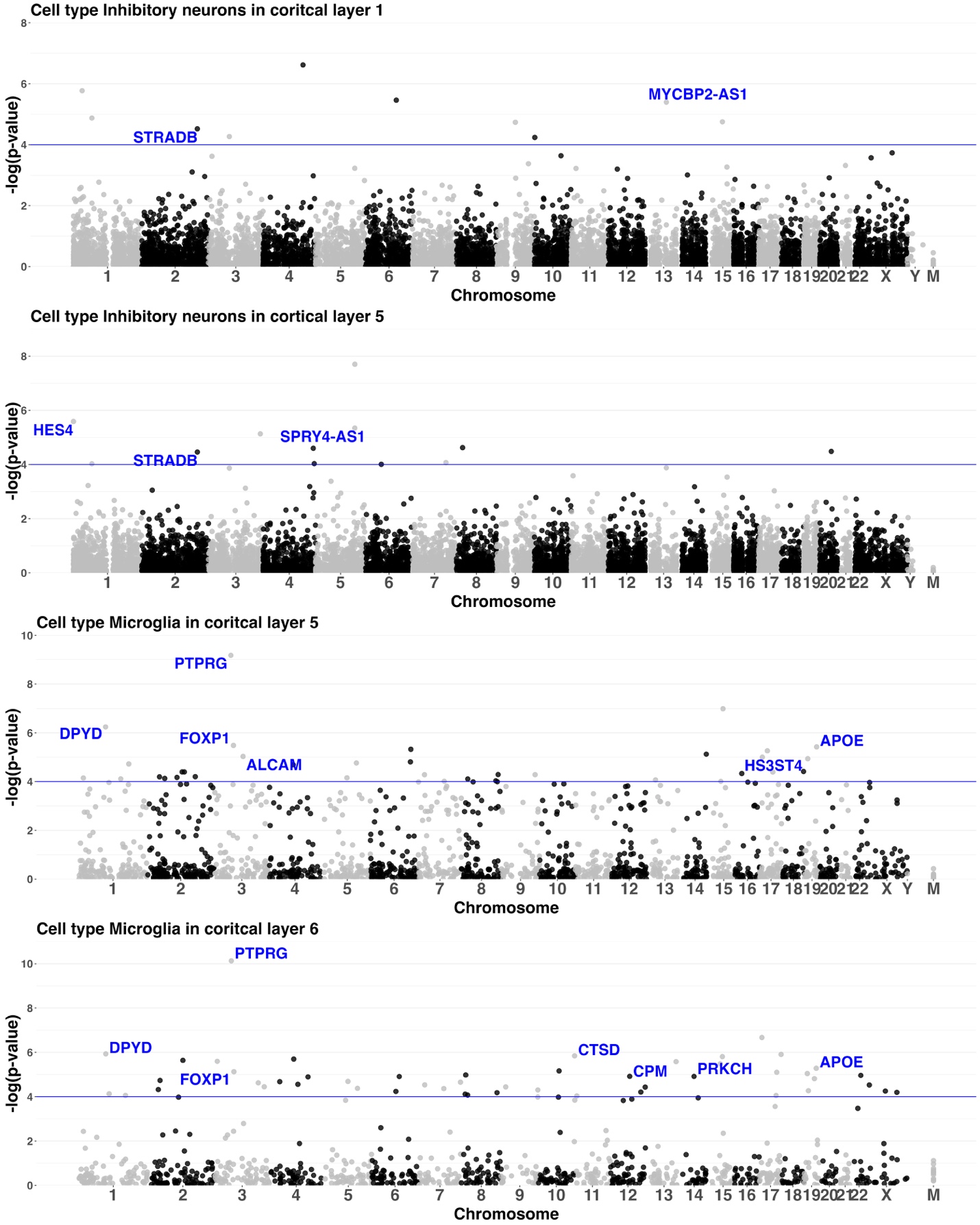

**Supp Fig 5**. **Manhattan plots of -log10(p-values) of the CLS-CTS DGE results of β-amyloid in Inhibitory neurons in layers 1 and 5, and Microglia in layers 5 and 6.** Genes labelled are also significant for either tangle density or/and cognitive decline.

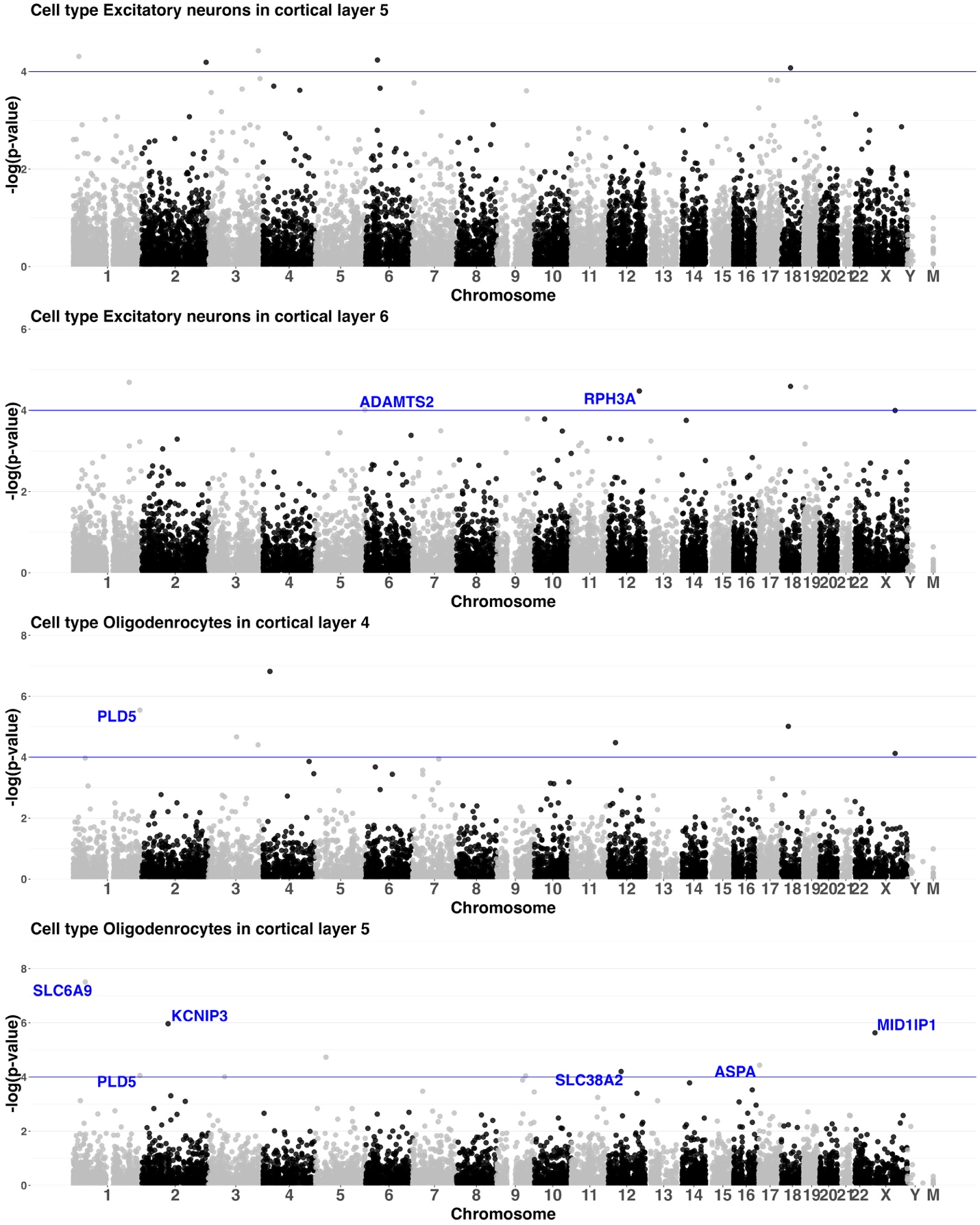

**Supp Fig 6. Manhattan plots of -log10(p-values) of the CLS-CTS DGE results of tangle density in Excitatory neurons in layers 5 and 6, and Oligodendrocytes in layers 4 and 5.** Genes labelled are also significant for either tangle density or/and cognitive decline.

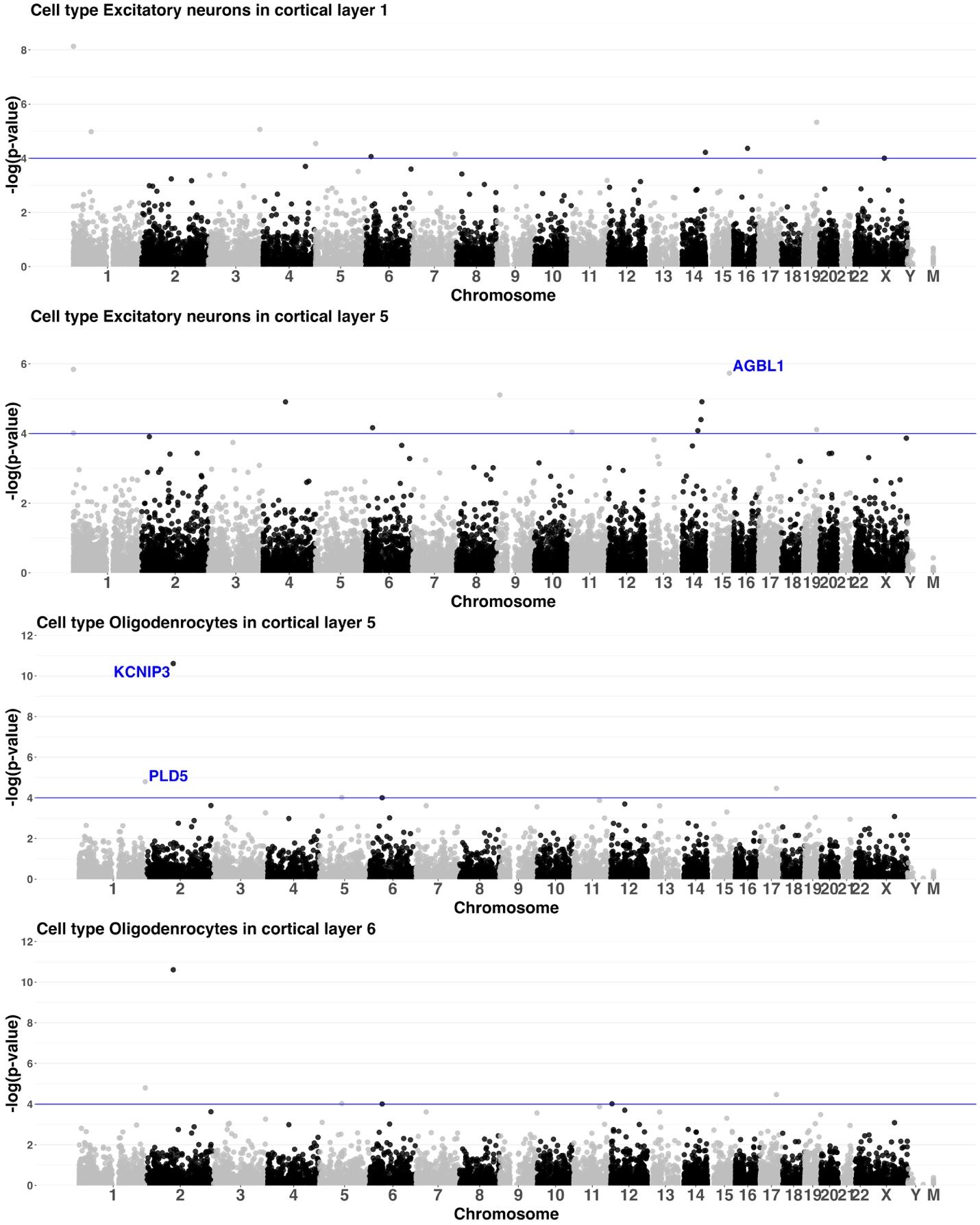

**Supp Fig 7.** **Manhattan plots of -log10(p-values) of the CLS-CTS DGE results of cognitive decline in Excitatory Neurons in layers 1 and 5, and Oligodendrocytes in layers 5 and 6.** Genes labelled are also significant for either tangle density or/and cognitive decline.

| **Reference ST Sample** | **Layer 1** | **Layer 2** | **Layer 3** | **Layer 4** | **Layer 5** | **Layer 6** | **WM** |
| --- | --- | --- | --- | --- | --- | --- | --- |
| **151673** | 129272  8.36% | 219182  14.17% | 403468  26.08% | 285185  18.44% | 217857  14.08% | 142103  9.19% | 149727  9.68% |
| **151674** | 148395  9.59% | 161342  10.43% | 226831  14.66% | 282293  18.25% | 294886  19.06% | 164303  10.62% | 268744  17.37% |
| **151675** | 181263  11.72% | 168230  10.88% | 225899  14.60% | 200518  12.96% | 339228  21.93% | 141716  9.16% | 289940  18.74% |
| **151676** | 153932  9.95% | 140889  9.11% | 202204  13.07% | 338649  21.89% | 297308  19.22% | 172607  11.16% | 241205  15.59% |
| **151507** | 298626  19.31% | 290513  18.78% | 441918  28.57% | 147273  9.52% | 124367  8.04% | 67245  4.35% | 176852  11.43% |
| **151508** | 333252  21.54% | 457270  29.56% | 338040  21.85% | 92885  6.01% | 97005  6.27% | 85110  5.50% | 143232  9.26% |
| **151509** | 488758  31.60% | 206956  13.38% | 250876  16.22% | 129976  8.40% | 146990  9.50% | 123642  7.99% | 199596  12.90% |
| **151510** | 583932  37.75% | 380507  24.60% | 243303  15.73% | 47180  3.05% | 28594  1.85% | 43634  2.82% | 219644  14.20% |
| **73,74,75,76** | 177324  11.46% | 203133  13.13% | 289453  18.71% | 199467  12.90% | 313961  20.30% | 238829  15.44% | 124627  8.06% |
| **73,74,09** | 238936  15.45% | 194827  12.60% | 394354  25.49% | 196243  12.69% | 237771  15.37% | 176705  11.42% | 107958  6.98% |
| **07,08,09,10** | 393851  25.46% | 333369  21.55% | 444032  28.71% | 90348  5.84% | 61191  3.96% | 54249  3.51% | 169754  10.97% |

**Supp Table 1. Numbers of cells in the ROS/MAP snRNA-seq data and their proportions in each cortical layer and WM that were inferred by CeLEry based on different reference ST data.** Reference ST samples 151673, 151674, 151675, 151676 are sections from one brain, while samples 151507, 151508, 151509, 151510 are sections from another brain. Multiple section samples from the same brain were combined as one reference “sample 73,74,75,76” and “sample 07,08,09,10”. Multiple section samples (151673, 151674, and 151509) from different brains were combined as one reference “sample 73,74,09”.

|  | **151676** | **151675** | **151674** | **151673** | **151507** | **151508** | **151509** | **151510** | **73,74,75,76** | **07,08,09,10** | **73,74,09** |
| --- | --- | --- | --- | --- | --- | --- | --- | --- | --- | --- | --- |
| **Layer 1** | 1.84 | 1.42 | 1.27 | 0.62 | 0.59 | 0.48 | 1.46 | **0.36** | 0.64 | 0.44 | 1.17 |
| **Layer 2** | 0.62 | 0.31 | 0.37 | 0.21 | 0.14 | 0.38 | 0.38 | 0.40 | **0.13** | 0.15 | 0.16 |
| **Layer 3** | 0.84 | 0.67 | 0.67 | **0.13** | 0.63 | 1.05 | 0.81 | 1.16 | 0.23 | 0.47 | 0.16 |
| **Layer 4** | 0.38 | 0.46 | 0.31 | **0.17** | 1.68 | 1.77 | 0.53 | 1.34 | **0.25** | 1.26 | 0.36 |
| **Layer 5** | 0.18 | 0.10 | 0.06 | 0.29 | 1.96 | 1.90 | 0.45 | 1.61 | 0.25 | 1.53 | 0.68 |
| **Layer 6** | 0.62 | 0.26 | **0.13** | 0.49 | 1.74 | 2.18 | 3.10 | 2.79 | 0.72 | 2.22 | 0.91 |
| **WM** | 0.96 | 0.50 | **0.43** | 0.62 | 1.60 | 2.35 | 2.28 | 2.01 | 0.94 | 1.53 | 0.94 |
| **Sum** | 5.43 | 3.72 | 3.25 | **2.54** | 8.34 | 10.11 | 9.02 | 9.67 | 3.16 | 7.60 | 4.40 |

**Supp Table 2. KL divergences between the “ground truth” CosMx ST data and inferred results for ROSMAP scRNA-seq data by using different LIBD reference ST data.** Reference ST samples 151673, 151674, 151675, 151676 are sections from one brain, while samples 151507, 151508, 151509, 151510 are sections from another brain. Multiple section samples from the same brain were combined as one reference “sample 73,74,75,76” and “sample 07,08,09,10”. Multiple section samples (151673, 151674, and 151509) from different brains were combined as one reference “sample 73,74,09”. A smaller KL divergence indicates greater similarity to the CosMx data, indicating more accurate spatial location inferences. The inferred results obtained by using sample 151673 showed the smallest KL divergence, which were used for follow-up DGE analyses.
